# Periodic dietary restriction of animal products induces metabolic reprogramming in humans with effects on health

**DOI:** 10.1101/2024.04.19.24306061

**Authors:** Konstantinos Rouskas, Ozvan Bocher, Alexandros Simistiras, Christina Emmanouil, Panagiotis Mantas, Anargyros Skoulakis, Y.C. Park, Alexandros Dimopoulos, Stavros Glentis, Gabi Kastenmüller, Eleftheria Zeggini, Antigone S. Dimas

**Author notes:** equal contribution (Konstantinos Rouskas, Ozvan Bocher).

## Abstract

**Objectives:** Dietary interventions can be a powerful tool for the prevention and treatment of diseases, but the molecular mechanisms through which diet affects health remain underexplored in humans. Generating robust evidence on the molecular impact of specific dietary patterns in humans will help us harness the power of dietary interventions to improve health.

**Methods:** Here, we compare plasma metabolomic and proteomic profiles between dietary states for a unique group of individuals (N=200) who alternate between omnivory and restriction of animal products for religious reasons. We also contrast findings to a control group of continuously omnivorous individuals (N=211).

**Results:** We find that short-term animal product restriction drives reductions in levels of lipid classes and of branched-chain amino acids, not detected in the control group, and results in metabolic profiles associated with decreased risk for all-cause mortality. We show that 23% of restriction-associated proteins are druggable targets and reveal that pro-longevity hormone FGF21 and seven additional proteins (FOLR2, SUMF2, HAVCR1, PLA2G1B, OXT, HPGDS, SPP1) display the greatest magnitude of change upon restriction. Through Mendelian randomization we demonstrate potentially causal effects of FGF21 and HAVCR1 on risk for type 2 diabetes, of HPGDS on BMI, and of OXT on risk for lacunar stroke.

**Conclusions:** We demonstrate that short-term restriction of animal products results in metabolic reprogramming with mostly positive effects on health and emphasise high-value targets for pharmacological intervention.

## 1. Introduction

Dietary interventions that involve the restriction of energy or of particular nutrients, without malnutrition, have been shown to delay aging and extend healthspan and lifespan in diverse species [1–5]. Although the type of restriction may vary (e.g. restriction of the level, type and timing of food consumption), studies have highlighted partially overlapping biological mechanisms of action, in evolutionarily conserved signalling pathways, as mediators of the effects of dietary restriction [1–5]. These pathways converge on key nodes of regulatory networks controlling aging-related processes and include mTOR, a protein a kinase acting in two complexes (mTORC1 and mTORC2) that mediates signalling in response to nutrient intake.

In humans, dietary restriction has been shown to have a protective effect against ageing-related disorders, including prevention of obesity and diabetes, cardiovascular disease, kidney disease, autoimmune and inflammatory conditions and cancer [2–4]. The last decades have seen an emergence of studies addressing the impact of patterns of dietary restriction on health, with plant-based diets in particular, having attracted a lot of interest [6–8]. Plant-based diets involve the restriction of animal product consumption, and practicing individuals typically present with lower BMI, lower serum LDL levels, and lower blood pressure [8]. Greater adherence to these diets has been linked to a reduced risk of cardiovascular disease, dyslipidemia, diabetes and certain types of cancer [8, 9], but also to some health risks, including lower bone mineral density and higher risk of stroke [6]. To date, few studies have addressed the molecular mechanisms underlying the effects of plant-based diets on health and disease, but work in this direction is emerging. Recently, a study comparing vegan versus ketogenic dietary interventions demonstrated effects on the innate and adaptive immune responses respectively [9]. Another study assessed plasma proteome differences in individuals from six dietary groups and revealed that individuals consuming plant-based diets displayed different profiles of proteins linked to gastrointestinal tract and kidney function [7]. A better understanding of the molecular effects of plant-based diets will enable us to harness their positive effects on health and to minimize possible detrimental consequences.

In this study, we have conducted molecular profiling for the first time, of a unique group of individuals from Greece who alternate between omnivory and restriction of animal products for religious reasons. The high consistency of the dietary pattern adhered to, and the predictability in switching between the two dietary states, means that this dietary behaviour shares characteristics with dietary intervention studies. To investigate the molecular impact of this dietary pattern, we compared plasma metabolomic and proteomic profiles of practicing individuals between dietary states, and to profiles obtained in parallel from a control group of continuously omnivorous individuals. We demonstrate extensive metabolic reprogramming upon animal product restriction and suggest that this dietary pattern can be harnessed to improve health and inform the development of interventions to prevent and treat disease, but a better grasp of underlying mechanisms is necessary to minimize possible negative effects.

## 2. Methods

### 2.1. FastBio population sample

The FastBio study was advertised during Autumn 2017 to the local communities of Thessaloniki, Greece through various means, including the press, radio, social media, and word of mouth. Candidate participants expressed their interest via telephone, email or via the project’s website (www.fastbio.gr). Following screening of over 1,000 candidates from the greater area of Thessaloniki through two interviews, 411 apparently healthy unrelated individuals, who met FastBio selection criteria (**Supplementary Text S1**) were included in the study. Participants belonged to one of two groups, specified by their diet: 200 individuals who followed a temporally structured dietary pattern of animal product restriction (periodically restricted, PR group) and 211 continuously omnivorous individuals who follow the diet of the general population and were not under any kind of special diet (non-restricted group, NR group).

PR individuals constitute a unique study group given their consistent adherence to a specific dietary pattern that involves alternating between omnivory and abstinence from meat, fish, dairy products and eggs (but not shellfish and molluscs) for 180-200 days annually (for detailed restriction pattern see **Supplementary Text S2, Figure 1b**). The PR diet is practiced for religious reasons as specified by the Greek Orthodox Church and is typically part of the culture of the family PR individuals are born into. It is initiated during childhood and is temporally structured with a specific periodicity involving four extended periods of restriction throughout the year, as well as restriction on Wednesdays and Fridays of each week. During periods of restriction, PR individuals also typically consume less alcohol. When not practicing restriction, PR individuals follow an omnivorous type of diet similar to the general population. Only PR individuals who had followed this dietary pattern for at least ten years were included in the FastBio study.

**Figure 1.**
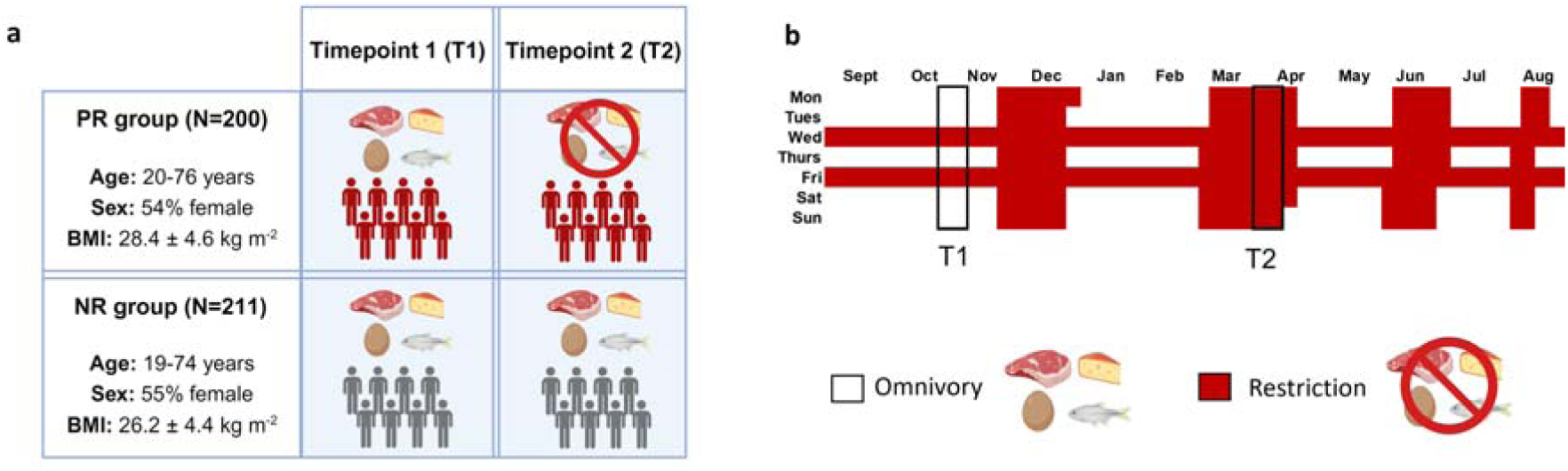
Study design and periods of animal product restriction. **a,** Schematic of study design. Periodically restricted (PR) individuals alternate between omnivory and animal product restriction for religious reasons. These two dietary states were profiled at timepoint 1 (T1) and timepoint 2 (T2) respectively. Non-restricted (NR) individuals are continuously omnivorous and were also profiled at T1 and T2. Created with BioRender.com. **b,** PR individuals practice animal product restriction for 180-200 days annually. Restriction is practiced during four extended periods throughout the year and on Wednesdays and Fridays of each week.

The FastBio population sample is described in ref[10], briefly, compared to NR individuals, PR were on average older (PR: 51.5 ± 13.5, NR: 45.0 ± 13.1), had higher BMI (PR: 28.4 ± 4.6, NR: 26.2 ± 4.4) and blood pressure (SBP PR: 127±19.0, NR: 121±19.7; DBP PR: 80±10.8, NR: 78±11.8) and were less likely to smoke (smokers PR: 6.5%, NR: 33.2%). Both groups consisted of slightly more female participants (PR: 54%, NR: 55%). Levels of physical activity were similar between groups and timepoints. The vast majority of participants were apparently healthy (according to self-reports) although a minority had underlying chronic conditions, including diabetes, arterial hypertension and hypothyroidism, for which treatment was being received. Very few participants were taking dietary supplements.

### 2.2. Collection of data, of biological material and measurement of traits

All FastBio participants were invited to two scheduled appointments at the Interbalkan Hospital of Thessaloniki. The first appointment took place in October-November 2017 (Timepoint 1 (T1)) and covered a period during which PR individuals has been on an omnivorous diet for 8-9 weeks (excluding Wednesdays and Fridays). The second appointment (Timepoint 2 (T2)) took place in March 2018 and was during Lent, covering a period where PR individuals had abstained from meat, fish, dairy products and eggs for at least three and a maximum of four weeks. A recall rate of 95% was achieved at T2, with 192 (out of 200) PR participants and 198 (out of 211) NR participants attending both appointments. For both timepoints, all appointments were scheduled between 7:30-9:30 am to minimize circadian effects, and were completed during a two-week window to minimize effects of seasonality for the specific sampling window. All participants gave their written informed consent and the study was approved by the local ethics committee (BSRC Alexander Fleming Bioethics Committee Approval 17/02/2017). For each participant, following overnight fasting, blood was drawn into EDTA-coated tubes and centrifuged to obtain plasma within 10-15 minutes of collection. Plasma was stored at -80□C. Aliquots were taken forward to generate metabolomics and proteomics data as described below.

### 2.3. Metabolomic profiling

For each sample, a volume of 100 μL was used for metabolomics analysis. Blood metabolite profiling of 249 metabolites was obtained using high-throughput NMR spectroscopy (Nightingale Health Plc, biomarker quantification library 2020). The panel is mostly composed of lipids and includes 168 absolute metabolite levels and 81 derived measures (ratios and percentages). Among the 801 samples, two could not be quantified, one due to a lack of material and one due to a technical error. Raw data corresponding to absolute concentrations or ratios of metabolites for the 799 remaining individuals were used for further analysis.

### 2.4. Proteomic profiling

For each sample, a volume of 40 µL was used for proteomics analysis. Plasma protein levels were measured on the Olink Explore 1536 platform (Olink Proteomics AB, Uppsala, Sweden) using the Neurology, Oncology, Cardiometabolic and Inflammation 384-plex panels (panel lot number B04414, B04412, B04413, B04411, respectively). Olink Explore is a protein biomarker platform that utilizes Proximity Extension Assay (PEA) technology with next-generation sequencing readout. Data generated from Olink are pre-processed and quality controlled using Olink data analysis software. Protein levels are reported in normalized protein expression values (NPX), which is a relative quantification unit in log_2_ scale with 1 NPX increase corresponding to doubled protein concentration.

### 2.5. Metabolite and protein quality control

All quality control (QC) steps are summarized in **Supplementary Figure S1**. QC procedures and statistical analyses were performed in R (version 4.1.3). Three individuals were removed from the analysis because they were first degree relatives with other participants. The success rate of metabolites and of samples were used to filter the data. The success rate was defined as the proportion of samples for which the metabolites had non-missing values. A threshold of 85% was applied for metabolites, resulting in the selection of 248 protein assays, and a threshold of 75% was applied for samples, resulting in the selection of 797 samples. The thresholds were based on the distribution of the QC metrics in the sample. The only metabolite removed was 3-Hydroxybutyrate (success rate 37%). Values below LOD were kept. Metabolite levels were log_2_-transformed prior to statistical analysis. We used a similar approach for proteins measured by the Olink Assay with the same threshold. This resulted in the selection of 1,464 protein assays and 793 samples. For quality assurance requirements, the panel includes three proteins that are quantified four times each (i.e. some proteins contain multiple IDs), the 1,464 protein assays correspond to 1,455 actual proteins. For simplicity, the term ‘proteins’ will refer to the Olink assays from now on. From the remaining data, all the values which have failed either Olink Assay QC or Olink Sample QC were set as missing while values below limit of detection (LOD) were kept in the downstream analyses. NPX values being already in the log_2_ scale, no further transformation of the data was applied prior to statistical analysis.

Principal Component Analysis (PCA) was used to check for sample outliers. Missing data were imputed by the mean prior to the PCA. The PCA on metabolite levels revealed 14 outliers (**Supplementary Figure S2a**) as samples with more than three standard deviations from the mean in the two first components (indicated by the red dashed lines). These samples were excluded from further analyses. On the PCA based on protein levels, a homogeneous group with no extreme outliers was observed (**Supplementary Figure S2b**). To maximize sample sizes and statistical power, no individual was removed on the basis of this PCA. The final dataset consisted of 777 and 793 samples for the metabolomics and proteomics statistical analyses respectively.

### 2.6. Statistical analysis

Differential abundance analysis for metabolite and protein levels was performed using the limma package (version 3.50.3) [11]. We used limma to record: A) changes in measured traits for each dietary group between timepoints (positive effects correspond to an increase at T2 compared to T1), and B) differences of measured traits between dietary groups for each timepoint (positive effects correspond to higher levels in PR compared to NR). To take into account the within-individual comparisons in the paired analyses, a linear mixed model was used where dietary groups and timepoints were treated as fixed effects and subject IDs were treated as random effects. All analyses were adjusted on sex, age squared, BMI and medication use (including medication for hypertension, thyroid, diabetes, osteoporosis and antiplatelets). Each medication variable was binary reporting whether individuals were taking this medication or not. It is to note that only a few participants are on medications (ranging from 7 individuals for the antiplatelets medication to 53 for the hypertension medication out of the 411 samples at T1). For the proteomics analysis, the mean value of the protein levels for each sample was included as an additional covariate. Limma was run separately on metabolite and protein levels and p-values were adjusted for multiple testing using the false discovery rate (FDR). Metabolites and proteins with an adjusted p-value lower than 5% were considered as differentially abundant.

### 2.7. Mortality score

To assess the effect of the metabolites on health, we used a mortality score developed by Deelen et al [12] combining levels of 14 selected metabolites from the Nightingale panel (XXL-VLDL-L, S-HDL-L, VLDL size, PUFA by FA ratio, glycine, lactate, histidine, isoleucine, leucine, valine, phenylalanine, acetoacetate, albumin and glycoprotein acetyls). Lower values of the score have been shown to be associated with a decreasing hazard ratio of overall mortality in a cross-sectional study, making it a surrogate for overall health of individuals. Following log_2_-transformation and scaling, the metabolic score was calculated for FastBio participants at both timepoints in both dietary groups. The scores were then regressed on the same covariates as in the differential expression analysis (age squared, sex, BMI, hypertension medication, diabetes medication, osteoporosis medication, cholesterol medication and antiplatelets medication) and compared between the groups using linear models.

### 2.8. Comparisons of animal product restriction-associated profiles to UK Biobank metabolite associations with complex diseases

To investigate the potential impact of metabolite changes linked to dietary restriction on metabolic health, we compared the pattern of metabolite associations observed in FastBio to the ones described by Julkunen et al [13] with multiple complex traits from the UK Biobank cohort (summary statistics downloaded from https://biomarker-atlas.nightingale.cloud/). We focused on ICD10 codes starting with ‘I-‘ and ‘E-‘, which correspond respectively to “diseases of the circulatory system” and to “endocrine, nutritional and metabolic diseases”, totalling 62 diseases. The t-statistics were computed from the estimates and standard errors, available in the downloaded file. We then performed PCA using the prcomp function in R on the computed t-statistics, also including the FastBio comparisons with the PR group (T2 vs T1) and at T2 (PR vs NR). We did not include the t-statistics of the comparisons within the NR group (T2 v T2) and at T1 (PR vs NR) as we detected only one and zero significant metabolites respectively. Additionally, we represented metabolite associations with each of the complex traits and in FastBio comparisons using a heatmap. For each metabolite class, we selected the most significant metabolite (as depicted in **Figure 2**).

**Figure 2.**
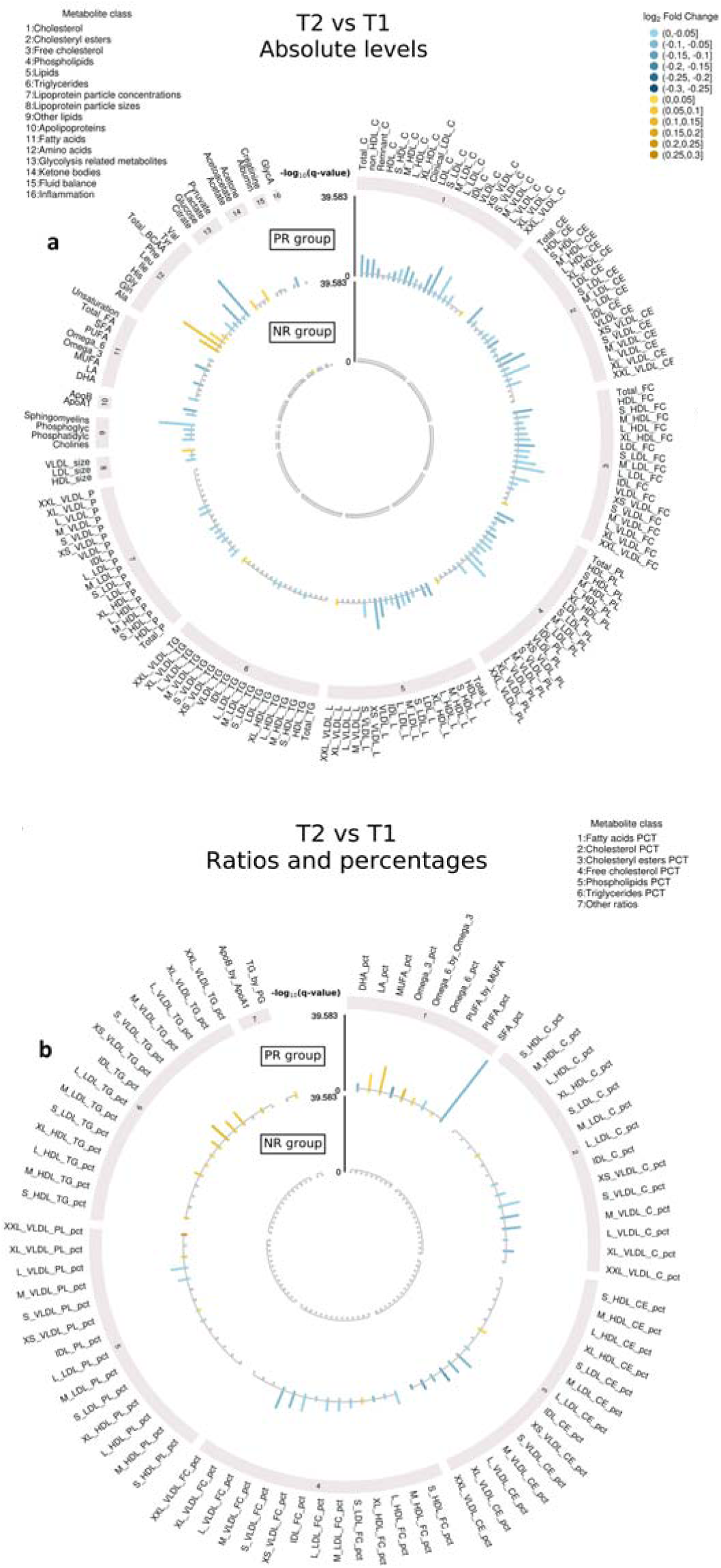
Differentially abundant metabolites detected from T1 to T2 for each dietary group. Changes in metabolite profiles are shown in the outer circle for PR individuals and in the inner circle for NR individuals (absolute values in panel **a**, ratios and percentages in panel **b**,). The -log_10_ of the FDR-adjusted p-value (q-value) is represented in the y-axis. Yellow bars represent an increase in metabolite levels from T1 to T2 whereas blue bars represent a decrease. Metabolites shown in grey are not significant.

### 2.9. Identifying proteins linked to FGF21 signalling

We investigated potential effects of FGF21 on proteins that were found to be differentially abundant between timepoints as a sensitivity analysis. For this, we repeated the differential protein expression analysis as described above, but restricted to proteins designated as differentially abundant (409, excluding FGF21 in the PR group; 210 in the NR group) in the primary analysis, and adjusted additionally for FGF21 levels. If a previously identified protein was no longer significant (FDR>0.05) after adjusting for FGF21 levels, we hypothesized that the two signals were not independent. We also conducted this sensitivity analysis between dietary groups (18 proteins tested at T1; 59 proteins tested at T2, after excluding FGF21). STRING DB (v 12.0) was queried to visualize interactions between FGF21 and proteins that did not retain their significance.

### 2.10. Animal product restriction-associated proteins as drug targets

We queried Open Targets [14] (March 8^th^ 2024) to investigate whether differentially abundant proteins unique to PR group are drug targets. For each differentially abundant protein that was a target for phase 1-4 drugs, we retrieved information on phase and status of the clinical trial, the approval status of the associated drugs, and their indications. Clinical trials with “Terminated”, “Withdrawn” or “Unknown” status were excluded. We focused on the analysis of phase 4 and approved drug-differentially abundant protein pairs and grouped their indications into broader Experimental Factor Ontology (EFO) disease categories to further explore their clinical translatability. Diseases categorized under the EFO terms ‘other disease’ or ‘other trait’ were further grouped, where possible, using the ancestral diseases as retrieved from Ontology Lookup Service (OLS) using the ‘rols’ R package [15].

### 2.11. Protein level heterogeneity analysis

To uncover proteins displaying the greatest magnitude of animal product restriction-associated change, we performed a heterogeneity analysis on the betas obtained from the limma association models. For this purpose, the Cochran’s statistics was used as follows:

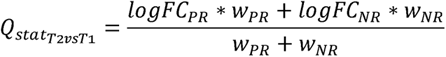

Where *logFc_PR_* corresponds to the log-fold change of the differential analysis between timepoints in the PR group (and in the NR group respectively), and *w_PR_* corresponds to the weight given to the analysis in the PR group which depends on the standard error *se_PR_* as:

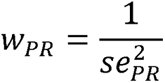

*w_NR_* is defined similarly for the NR group.

This statistic follows a chi-square distribution with one degree of freedom. To assess the statistical significance of the test, we used a Bonferroni-corrected significance threshold. We computed the effective number of proteins based on their correlation pattern using the procedure described by Li and Ji [16], giving a total of 1,432 independent tests. We then divided the classical 5% threshold by this value, resulting in a significance threshold of 3.5e- 5.

### 2.12. Animal product restriction-associated proteins in clinical trials

We queried https://clinicaltrials.gov/ (Oct 30^th^ 2023) for the top 23 most heterogenous differentially abundant proteins (FDR-corrected results from the heterogeneity analysis) inputting the protein name in the intervention/treatment field and reporting all clinical trials on record (phases 1-4).

### 2.13. Causal effects of proteins associated with animal product restriction on complex traits

To investigate the causal effect of the eight proteins identified in the heterogeneity analysis on age-related traits, we performed two-sample Mendelian randomization (MR). We focused on five complex traits: T2D, body mass index (BMI), coronary artery disease (CAD), lacunar stroke and essential hypertension. We selected the instrumental variables (IVs) for the eight proteins by using the protein quantitative trait locus (pQTLs) described in the UK Biobank [17], available in https://www.synapse.org/#!Synapse:syn51364943/files/. To match the ancestry of the FastBio cohort, we used the summary statistics from the European population. We focused on cis-pQTLs by defining windows at +/- 500kb from the genomic positions of the corresponding genes, obtained through GeneCards (**Supplementary Table S1**). We then performed clumping by using the 1000Genomes European reference panel, a r2 threshold of 0.001 in 10Mb windows, and a p-value threshold of 1e-5. The F-statistic for the remaining variants was computed as 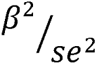 , and only variants with an F-statistic greater than 10 were considered as IVs. The summary statistics for the five complex traits were downloaded from publicly available databases, by minimizing sample overlap with the UK Biobank (**Supplementary Table S1**). We then performed two sample MR by using the package

TwoSampleMR [18]. We considered the inverse variance weighted (IVW) method, or the Wald ratio if only one IV was present, to assess statistical significance at a threshold of 1.61e-3, corrected for the 31 tests performed. To assess robustness of the results against the MR assumptions, we verified the direction of effect obtained using the weighted median and MR-Egger methods. Reported significant signals correspond to signals with a significant p-value and a concordant direction of effect with the sensitivity analyses. If heterogeneity was present (nominally significant p-value), we further applied MR-Presso [19] and considered the corresponding p-value after removing outliers.

## 3. Results

### 3.1. Population sample description and study design

Following screening of over 1,000 candidate participants, we recruited 411 apparently healthy individuals from Thessaloniki, Greece for the FastBio (Religious <u>Fast</u>ing <u>Bio</u>logy) study (**Figure 1a, Supplementary Text S1**). Participants belonged to one of two dietary groups: individuals who had undergone periodic restriction of animal products for religious reasons, for at least ten years, and individuals who were continuously omnivorous. Periodically restricted (PR) individuals (N=200, ages 20-76 years) voluntarily alternate between restriction and omnivory following the dietary regimen of the Greek Orthodox Church (**Supplementary Text S2**). This involves abstinence from meat, fish, dairy products and eggs (but not molluscs and shellfish) for 180-200 days annually, in a highly consistent and predictable manner. Restriction is practiced during four extended periods throughout the year, as well as on Wednesdays and Fridays of each week (**Figure 1b**). Non-restricted (NR), control individuals (N=211, ages 19-74 years) were continuously omnivorous and did not practice any type of dietary restriction. Participants were profiled at two timepoints: T1 in autumn, covering a period of omnivory for both dietary groups, and T2 in early spring, covering a three-to-four-week period of restriction for PR individuals, during Lent (**Figure 1b**). Typically, during this period, PR individuals undergo restriction of protein (as a proportion of total energy) which is not accompanied by a decrease in total energy intake [20, 21].

### 3.2. Animal product restriction is associated with a prominent shift in metabolomic profiles

We first sought to investigate how restriction of animal products affects plasma metabolite levels through quantification of 249 metabolites. Following quality control (QC) (**Supplementary Figure S1, Supplementary Figure S2**), we analysed 248 metabolites from 777 samples (193 PR at T1 and 190 PR at T2; 202 NR at T1 and 192 NR at T2). We found a prominent shift in metabolite profiles for the PR group with over half (N=168) of quantified metabolites detected at altered levels upon restriction (**Figure 2, Supplementary Table S2**). In the NR group, only pyruvate was detected at slightly altered levels (**Figure 2, Supplementary Table S2**). Most changes associated with animal product restriction (134 out of 168) involved decreased metabolite levels with reductions in multiple lipid classes including cholesterol, cholesterol esters, free cholesterol, phospholipids, and lipids from most lipoprotein particle types (HDL, IDL, LDL) and sizes. Decreased levels were also found for sphingomyelins, for the ratio of saturated to total fatty acids (SFA %) and for both ApoA1 and ApoB. Lower levels of ApoB, which is a major constituent of LDL-related lipids, are associated with decreased risk of cardiovascular events [22], while HDL-related lipids, and its major constituent ApoA1, both have cardioprotective effects [23, 24]. Reductions in levels of LDL-related lipids were of a greater magnitude compared to those recorded for HDL-related lipids. Similar patterns have been revealed in studies of individuals following a vegan diet [25, 26], a dietary pattern in close proximity to animal product restriction, and likely reflect the overall lower intake of animal fat. Furthermore, although we did not detect changes in triglyceride levels, we identify a striking pattern of increased proportions of triglycerides in VLDL particles (**Figure 2b**), pointing towards an increased rate of triglyceride removal from the blood to the liver.

We also found altered levels for eight out of the nine amino acids quantified upon restriction (**Figure 2a**), with a ∼7% decrease detected for absolute concentrations of total BCAAs, driven by decreases in valine (9%) and leucine (5%), but not isoleucine. BCAAs, which are strong agonists of mTORC1, make up ∼20% of amino acids in meat, fish, eggs and nuts [27] and elevated levels in humans have been linked to poor health outcomes including obesity, type 2 diabetes and cardiovascular disease [28–30]. BCAA restriction on the other hand has been linked to reduced insulin resistance [31] and to increased levels of potent metabolic regulator and pro-longevity hormone FGF21 in some [32], but not all studies [33]. Furthermore, although protein restriction has been linked to decreased fasting blood glucose levels in overweight or obese males [34], we did not find changes in glucose levels in the present study, which comprises participants from both sexes who span the range of BMI.

Given the similarity of the dietary pattern under study to a vegan diet, we compared our findings to those from a cross-sectional study investigating profiles of the same 249 metabolites in vegans and meat-eaters [35], and replicated 82% of reported associations (**Supplementary Figure S3**). This suggests that a short course of animal product restriction results in a metabolite profile that shares similarity to profiles of permanent vegans. Additionally, we found 58 previously unreported associations for metabolites including small VLDL, medium LDL, and large HDL classes, with decreases in cholesterol-associated levels upon restriction and identified 27 new associations for metabolites not analysed in the above study, including total BCAAs, pyruvate and LDL- and medium HDL-related lipids (**Supplementary Figure S3**).

We also examined differences of metabolite profiles between dietary groups for each timepoint and found that when both groups are omnivorous, they share identical profiles (**Supplementary Text S3, Supplementary Figure S4**). Upon dietary restriction however we detected differences in the levels of 102 metabolites, of which 97 (95%) were also detected in the PR group from T1 to T2, supporting the idea that these changes are directly linked to animal product restriction. Our findings suggest that metabolomic reprogramming is rapid, but also likely transient.

### 3.3. Animal product restriction results in metabolomic profiles associated with reduced risk for all-cause mortality and with beneficial effects against complex diseases

To investigate the impact of animal product restriction-associated metabolite profiles on health, we applied a 14-metabolite score that reflects risk for all-cause mortality, with lower scores associated with a lower mortality risk [12]. Nine of the 14 metabolites examined were detected at altered levels upon dietary restriction (VLDL size, PUFA by MUFA ratio, histidine, leucine, valine, phenylalanine, acetoacetate, XXL_VLDL_L, S_HDL_L). Animal product restriction led to a significant decrease in the mortality score (mean score 0.11 at T1 vs -0.91 at T2, p = 0.011), that was not found in the control group (mean score 0.079 at T1 vs 0.70 at T2, p = 0.431) (**Supplementary Figure S5**). To further address the impact of restriction of animal products, we compared restriction-associated changes to associations between metabolite levels and age-associated traits from the UK Biobank [13]. Focusing on “diseases of the circulatory system” and “endocrine, nutritional and metabolic diseases”, metabolite profiles linked to animal product restriction mapped away from metabolite profiles associated with increased risk of cardiovascular outcomes (including acute myocardial infarction and chronic ischemic heart disease), diabetes, obesity, and dyslipidemia (**Supplementary Figure S6a**). Differences between animal product restriction-associated metabolite profiles and disease-associated profiles were driven chiefly by differences in lipid-related classes, ketone bodies and fatty acid percentages (**Supplementary Figure S6b)**.

### 3.4. Animal product restriction is associated with changes in the levels of multiple plasma proteins

We next sought to investigate how animal product restriction affects plasma protein levels. We quantified levels for 1,472 proteins and following QC (**Supplementary Figure S1, Supplementary Figure S2)**, we report results for 1,464 proteins from 793 individuals (199 PR at T1 and 191 PR at T2; 208 NR at T1 and 195 NR at T2). We detected 410 and 201 proteins whose levels changed from T1 to T2 in the PR and NR groups respectively (**Figure 3, Supplementary Table S2**). Protein level changes unique to each group were almost five-fold higher in PR (264) compared to NR individuals (55), with increased levels of FGF21 being the most prominent change recorded, and unique to the PR group. In addition to PR- unique changes, we found that 146 proteins changed levels in both groups, with the same direction of effect, some of which likely capture seasonal effects. For example, MNDA and ANXA3, proteins associated with neutrophils displayed decreased levels at T2 in both groups. We [10] and others [36] have shown that neutrophils, display strong seasonal effects, with their numbers decreasing during spring. Changes unique to the NR group were of lower effect size and significance and may also represent seasonal changes that are suppressed in PR individuals through dietary restriction. For example, the most significantly altered NR- unique protein, PPP1R9B is a substrate for GSK3β, a crucial circadian clock regulator [37] while PCSK9, a target for LDL-lowering drugs, displays diurnal variation, with levels of daylight duration likely driving seasonal effects [38].

**Figure 3.**
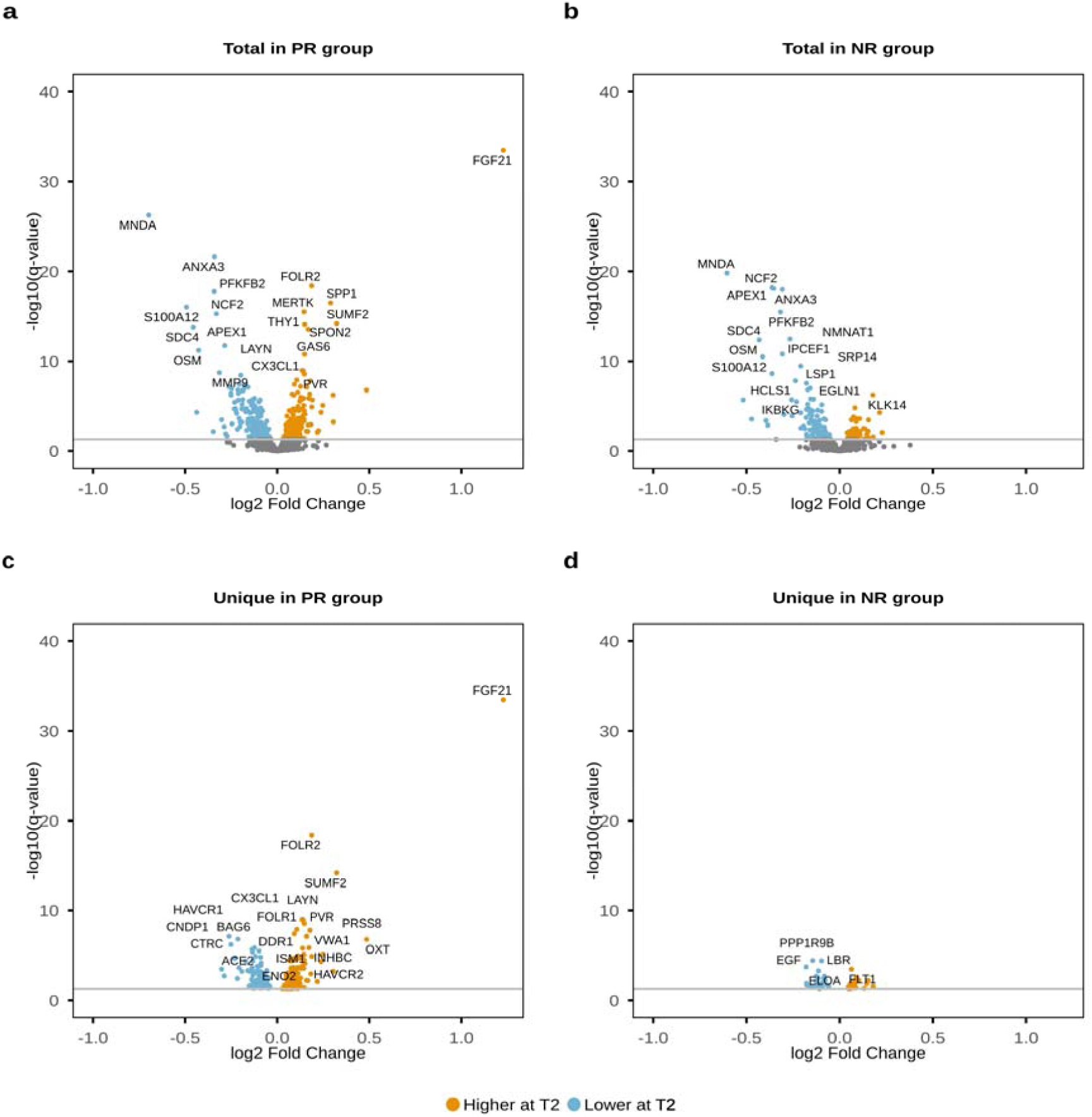
Differentially abundant proteins detected from T1 to T2 for each dietary group. Total differentially abundant proteins detected in the PR group (**a**) and in the NR group (**b**). Unique differentially abundant proteins detected in the PR group (**c**) and in the NR group (**d**). Proteins with increased levels at T2 are shown in yellow whereas proteins with decreased levels are shown in blue. Proteins shown in grey are not significant.

We also examined differences in protein abundance between dietary groups and found that 18 and 60 proteins displayed different levels at T1 and T2 respectively (**Supplementary Text S3, Supplementary Figure S7**). Of these, 10 proteins were consistently different between groups, likely reflecting long-term effects of animal product restriction. The most prominent example is ALPP, that was detected at lower levels in the PR group at both timepoints. Plasma levels of this enzyme are thought to derive from neutrophils, which have been found to be consistently lower in the PR group at T1 and T2 [10].

We next explored links between metabolites and proteins using unsupervised correlation analysis, we detected two main correlation modules. The first component captured animal product restriction-associated effects, with five PR-unique differentially abundant proteins (HAVCR1, ESM1, SPP1, SPON2, FGF21) correlating with IDL particles. The second component captured effects not linked to animal product restriction with five proteins (LDLR, NPY, AGRP, CD38, MFGE8) correlating positively with L and XL VLDL particles and with triglycerides (**Supplementary Text S4, Supplementary Figure S8**).

### 3.5. Proteins associated with animal product restriction are druggable targets

Given that interactions between drugs and their targets affect drug action, we sought to determine how many of the proteins whose abundance is altered following animal product restriction (N=264) are targets for known drugs. We found that 62 (23%) of proteins are targets for phase 1-4 drugs, and of these 28 (11%) are targets for phase 4, approved drugs (**Figure 4**). Indications covered by the above drugs include mostly conditions associated with aging such as cancer, cardiovascular, immune system and inflammatory, and visual system diseases (**Figure 4, Supplementary Table S3**). Notably, over half of the druggable proteins were found to be targets for multiple drugs, with EGFR for example being targeted by 56 phase 1-4 drugs, of which 20 are approved therapeutics for the treatment of various cancer types (**Figure 4, Supplementary Figure S9, Supplementary Table S3**).

**Figure 4.**
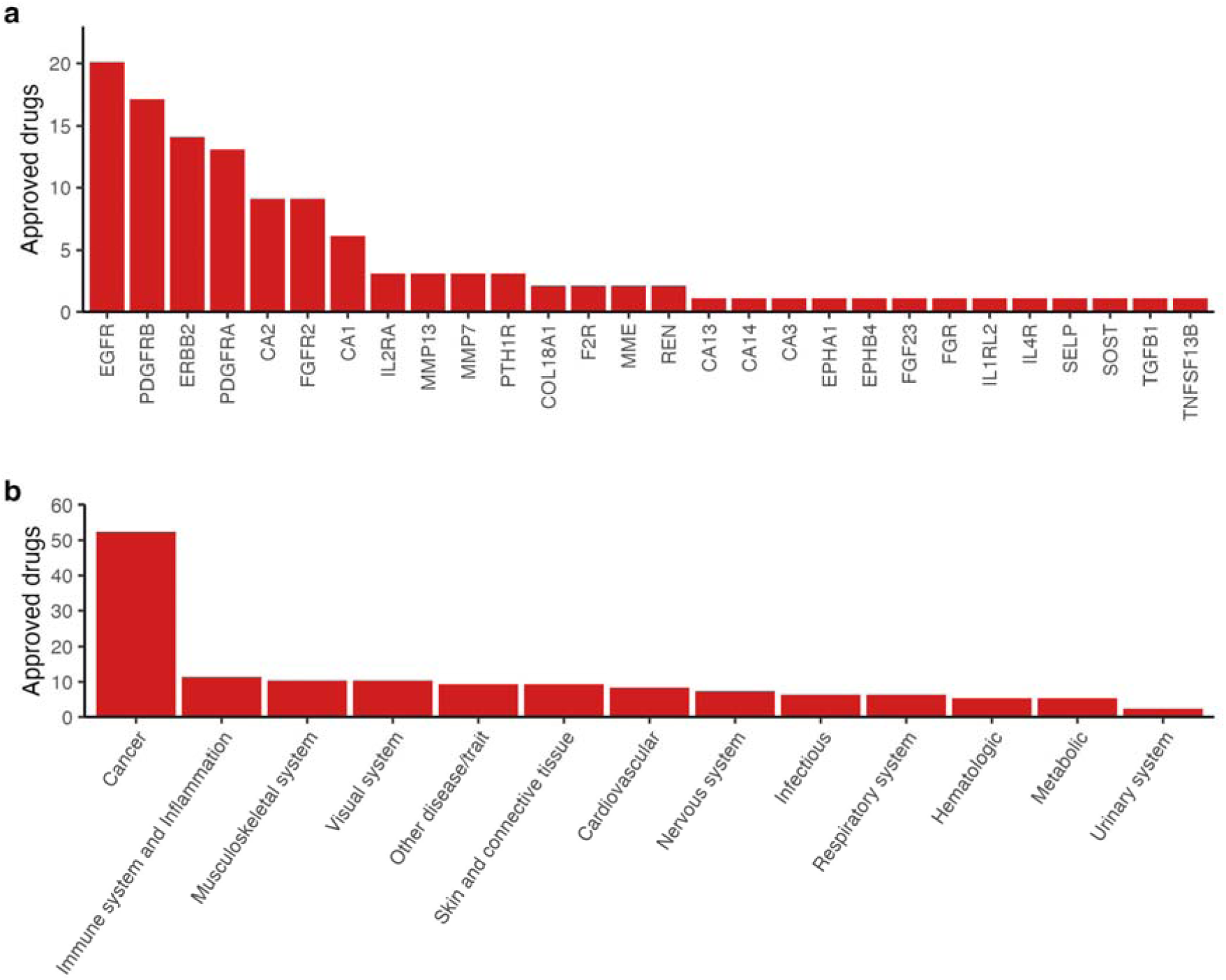
PR-unique differentially abundant proteins as targets of approved drugs and associated disease indications. **a**, Number of approved drugs (excluding withdrawn drugs) that target differentially abundant proteins detected only in the PR group. **b**, Number of approved drugs (from above) per disease indication.

### 3.6. Multiple proteins displaying the greatest magnitude of change upon animal product restriction are being evaluated in clinical trials

Due to pre-enrichment for specific biological functions of the panel of assayed proteins, we were underpowered to uncover over-represented pathways for proteins detected at altered levels from T1 to T2 (**Supplementary Text S5**). To focus on proteins most affected by restriction of animal products, we conducted a heterogeneity analysis that aimed to identify proteins for which the effect size of change from T1 to T2 was significantly different between the two dietary groups. Following correction for 1,432 independent tests, we highlight eight proteins most affected by animal product restriction: FGF21, FOLR2, SUMF2, HAVCR1, PLA2G1B, OXT, HPGDS, and SPP1 **(Figure 5**, **Table 1)**. Of these, FGF21, FOLR2 and HAVCR1 were detected at significantly different levels in permanent vegetarian and vegan individuals compared to meat-eaters, in a recent study [7], with differences being in the same direction as the effects reported here. Using a more permissive 5% FDR correction, we identify 15 additional proteins and underline that over half of these (**Table 1**) are currently being evaluated in clinical trials for their pharmacological value for age-related outcomes including type 2 diabetes, obesity, cardiovascular disease, cancer, and renal function (**Table 1**).

**Figure 5.**
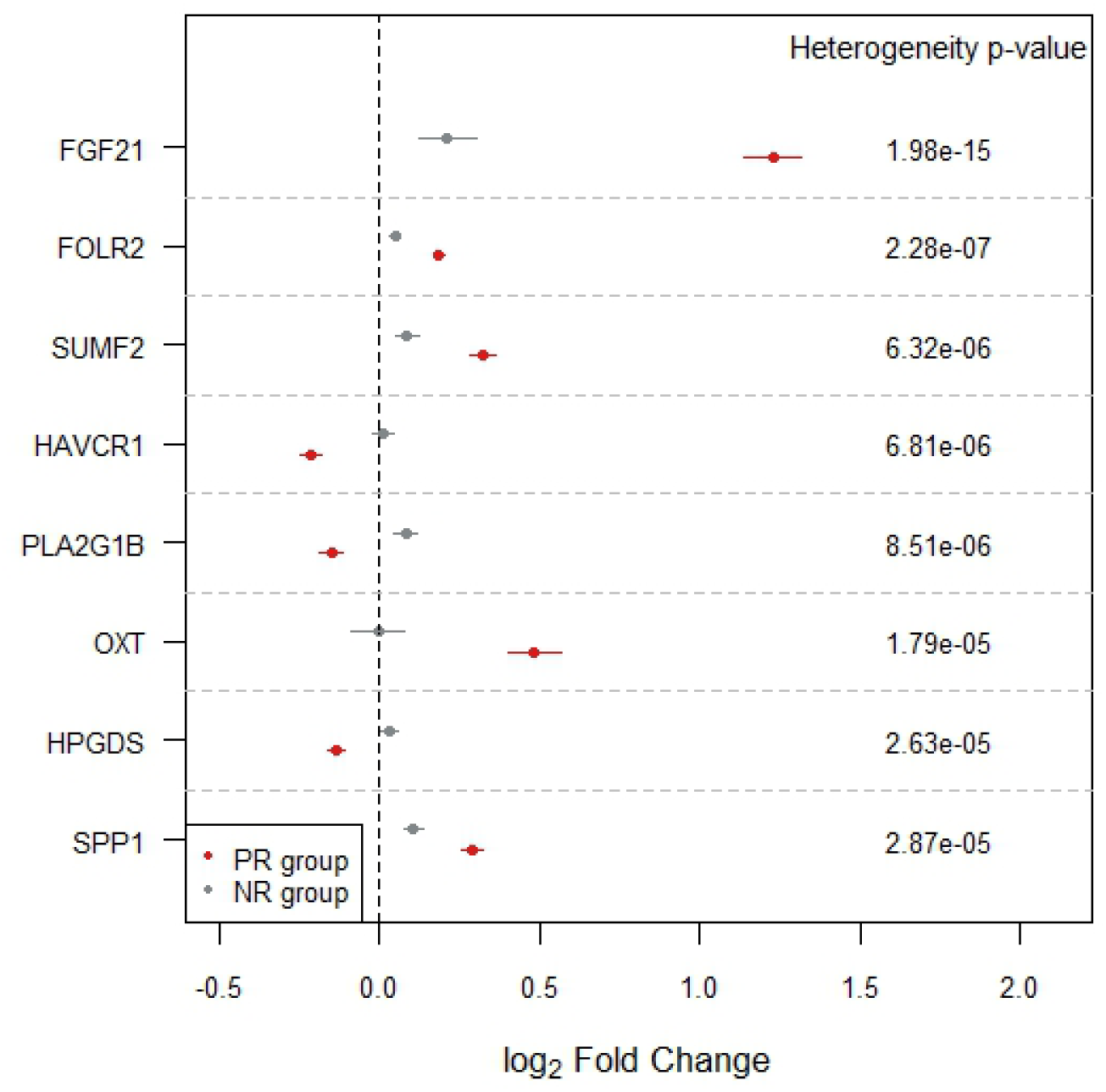
Proteins displaying the greatest magnitude of change linked to animal product restriction. Eight proteins showing the greatest effect size changes from T1 to T2 and between dietary groups were identified through heterogeneity analysis using the Cochran’s statistics (associated p-value indicated in ‘Heterogeneity p-value’). Associations between levels of each protein and timepoints are shown in red for the PR group and in grey for the NR group.

**Table 1.** FDR-corrected proteins displaying the greatest magnitude of change upon animal product restriction and associated clinical trials.

| **Protein** | **Change** | **No. trials** | **Indicative diseases and conditions** |
| --- | --- | --- | --- |
| **FGF21** | ↑ | 39 | gestational and type 2 diabetes, insulin resistance, obesity, liver health |
| **FOLR2** | ↑ | 0 |  |
| **SUMF2** | ↑ | 0 |  |
| **HAVCR1** | ↓ | 24 | renal function |
| **PLA2G1B** | ↓ | 54 | atherosclerosis, coronary disease, renal function, type 2 diabetes |
| **OXT** | ↑ | 773 | pregnancy and labour, infertility, bleeding, obesity, neurological disorders, substance dependence, pain, social cognition |
| **HPGDS** | ↓ | 1 | pulmonary health |
| **SPP1** | ↑ | 25 | cancer, infant growth and development |
| **CNDP1** | ↓ | 0 |  |
| **PRSS8** | ↑ | 0 |  |
| **BAG6** | ↓ | 1 | hemoglobin sickle cell disease |
| **FGF23** | ↓ | 108 | renal function, cardiovascular health, metabolic health, X-linked hypophosphatemia |
| **ASAH2** | ↓ | 0 |  |
| **LRIG1** | ↓ | 0 |  |
| **PVR** | ↑ | 0 |  |
| **THY1** | ↑ | 0 |  |
| **FOLR1** | ↑ | 58 | fallopian tube, ovarian, breast, cervical and peritoneal cancer |
| **CTRC** | ↓ | 0 |  |
| **DDR1** | ↑ | 2 | advanced cancer |
| **FST** | ↓ | 7 | cardiovascular health, muscular dystrophy, infertility, cancer |
| **FGFR2** | ↑ | 57 | gastric and bile duct cancer |
| **ACE2** | ↓ | 55 | COVID-19, cardiovascular health, renal function |
| **MVK** | ↑ | 5 | mevalonate kinase deficiency, Behcet's disease |

Change direction refers to the change in protein abundance from timepoint 1 to timepoint 2 for the PR group.

### 3.7. Pro-longevity hormone and potent metabolic regulator FGF21 is the protein most affected by animal product restriction

Our most prominent finding associated with animal product restriction was an increase in levels of liver-derived hormone FGF21. In humans and mice, FGF21 has key roles in regulation of energy homeostasis, lipid and glucose metabolism, and insulin sensitivity[2, 39, 40]. Moreover in mice, prolonged overexpression of this hormone has been shown to extend lifespan [41]. FGF21 is induced by dietary protein restriction [42] and by restriction of specific amino acids [2] and mediates its effects directly on adipose tissue and through signalling to the brain to regulate macronutrient preference [43]. Currently, it is in clinical trials for conditions including type 2 diabetes, insulin resistance, obesity, and non-alcoholic steatohepatitis (**Supplementary Table S4**). To date, potent beneficial effects on lipid profiles have been reported, but the end points of glycemic control have not been met, and the impact of FGF21 on glucose homeostasis in humans is not yet clear [44].

FGF21 signalling in humans therefore remains to be elucidated and its complexity may be exacerbated by the existence of FGF21 inactivation enzymes [39] such as FAP. We found that upon animal product restriction FAP levels decreased, potentially resulting in extension of the short half-life of FGF21 (0.5–1.5 hours). To address the complexity of FGF21 signalling more broadly, we conducted differential abundance analysis on the 409 proteins associated with animal product restriction (excluding FGF21), and adjusted additionally on FGF21 levels. Results did not change for the NR group, but in the PR group, 53 proteins were no longer significant, suggesting association of their abundance with FGF21 (**Supplementary Table S5**). Evidence for links to FGF21 exists for some of these proteins (e.g. FGF23, EGFR and CCL2) (**Supplementary Figure S10**), but for others, their connection to FGF21 remains unknown, rendering them good candidates for functional evaluation.

### 3.8. Animal product restriction is associated with increased abundance of proteins with key metabolic functions

In addition to FGF21, increased levels upon restriction were recorded for FOLR2, SUMF2 and OXT. FOLR2 imports folate to the interior of cells and together with vitamin B12, a micronutrient found exclusively in animal products, is an essential compound of the folate and methionine cycles [45]. Deficiency of vitamin B12, typical in vegan individuals [6], can mimic the effects of folate deficiency. Given that under conditions of folate deficiency, folate receptors are upregulated [46], we suggest that restriction-associated increased FOLR2 levels may be in part due to decreased vitamin B12 intake. Folate deficiency is also associated with high levels of homocysteine, typically observed in individuals consuming plant-based diets [4]. Homocysteinemia has been shown to accelerate processes driving aging [47] and is a risk factor for disorders including neurodegenerative and cardiovascular diseases, including stroke. SUMF2 is an inhibitor of the enhancing effects of SUMF1 on sulfatases [48] and causal effects on decreased levels of HDL, LDL and total cholesterol have been found [49]. Restriction of animal products therefore has likely inhibitory effects on sulfatase activity through increased levels of SUMF2, and may contribute to effects on the above lipids. OXT is a hypothalamic hormone known for its roles in parturition, lactation, social bonding, but also acts as a sensor of nutrient status [50]. In humans, decreased circulating OXT levels have been recorded in individuals with obesity or diabetes, and OXT administration is currently being explored as a treatment for obesity-related comorbidities [51] (**Supplementary Table S4**). Similarly to FGF21 [43], OXT shapes macronutrient preference and affects weight control [50].

SPP1 (OPN) was detected at increased levels in both dietary groups from T1 to T2, but the magnitude of change was greater for PR individuals. SPP1 has diverse functions including bone calcification, cell survival, proliferation and migration, and immune function [52] and is in clinical trials for indications linked to cancer (**Table 1**). In mice, overexpression of SPP1 in the retina and optic nerve slows visual system aging and restores vision in models of optic nerve damage [53]. SPP1 expression is induced by calcitriol, the hormonal metabolite of vitamin D, that is produced under low levels of calcium [52, 54]. Given the impact of sunlight on vitamin D, increased levels of SPP1observed for both dietary groups from T1 to T2, may be linked to seasonal effects. Furthermore, individuals practicing restriction of animal products typically have lower calcium intake [21], as do vegans, who display consistently lower levels of vitamin D [6]. The greater magnitude of change in SPP1 levels in the PR group may therefore be due to a combination of seasonal and dietary restriction-related effects.

### 3.9. Animal product restriction is associated with decreased abundance of proteins with immunometabolic functions

Decreased levels upon restriction were detected for HAVCR1, PLA2G1B and HPGDS. HACVR1 is a transmembrane receptor with roles in regulation of immune cell activity and renal regeneration [55]. It is abnormally expressed in a range of tumours, upregulated during renal injury [55], and is currently in multiple clinical trials mostly for indications linked to renal function (**Supplementary Table S4**). Recent work suggests that in addition to renal injury and cancer, the predictive value of this protein extends to multiple aging-associated diseases including disorders of the endocrine, nervous, circulatory and respiratory systems [56]. In rat primary renal tubular epithelial cells, administration of the saturated fatty acid (SFA) palmitate results in upregulation of HAVCR1 [57]. Decreased HAVCR1 levels are therefore consistent with the prominent reduction of SFA% detected upon restriction of animal products, reflecting the lower intake of saturated fats [35]. PLA2G1B is a secreted phospholipase A2 produced by pancreatic acinar cells [58], and is currently being investigated in clinical trials for a range of cardiometabolic conditions (**Supplementary Table S4**). *Pla2g1b* knock-out mice display reduced phospholipid digestion and concurrent attenuation of diet-induced obesity, insulin resistance, and atherosclerosis. Conversely, pancreatic acinar cell-specific overexpression of Pla2g1b has been linked to increased phospholipid digestion and to exacerbation of diet-induced obesity and insulin resistance [58]. Notably, members of the phospholipase A2 family have been linked to healthspan and longevity in humans in a study showing that PLA2G7 downregulation in adipose tissue mediated the beneficial effects on health of prolonged caloric restriction [26]. Adding to the idea that PLA2 protein family members influence pathways regulating longevity, genetic variants in pla2g10 in rockfish, a clade including 107 species that can live between 11 to 205 years, have been linked to lifespan [59]. Finally, HPGDS is an enzyme that catalyses the conversion of PGH2 to PGD2. PGD2 is a prostaglandin that acts as an early-phase mediator of inflammation [60] and augments disease activity, for example in asthma [61]. Decreased levels may therefore contribute to a tempering effect on inflammation and the role of HPGDS in pulmonary health is currently being evaluated in clinical trials (**Supplementary Table S4**).

### 3.10. Proteins affected by restriction of animal products have potentially causal effects on complex diseases

Given the key immunometabolic roles of the eight proteins above, we sought to investigate causal effects of their genetically regulated levels on complex traits using Mendelian randomization (MR). We focused on traits and diseases from the UK Biobank with associated metabolite profiles that mapped the furthest away from metabolite profiles of PR individuals (**Supplementary Figure S6**), namely type 2 diabetes, body mass index (BMI, proxying obesity), coronary artery disease, lacunar stroke (proxying atherosclerosis) and essential hypertension. At the Bonferroni corrected significance threshold, we identified a protective effect of FGF21 and HACVR1 on T2D risk and a causal effect of HPGDS on decreased BMI (**Figure 6, Supplementary Table S1**). We also found that increased OXT levels drive an increased risk for lacunar stroke, which is in the opposite direction of the suggested protective effects of OXT against cardiovascular risk in the literature [62]. However, vegetarian individuals from the EPIC-Oxford study display an increased risk of stroke compared to meat-eating individuals [6].

**Figure 6.**
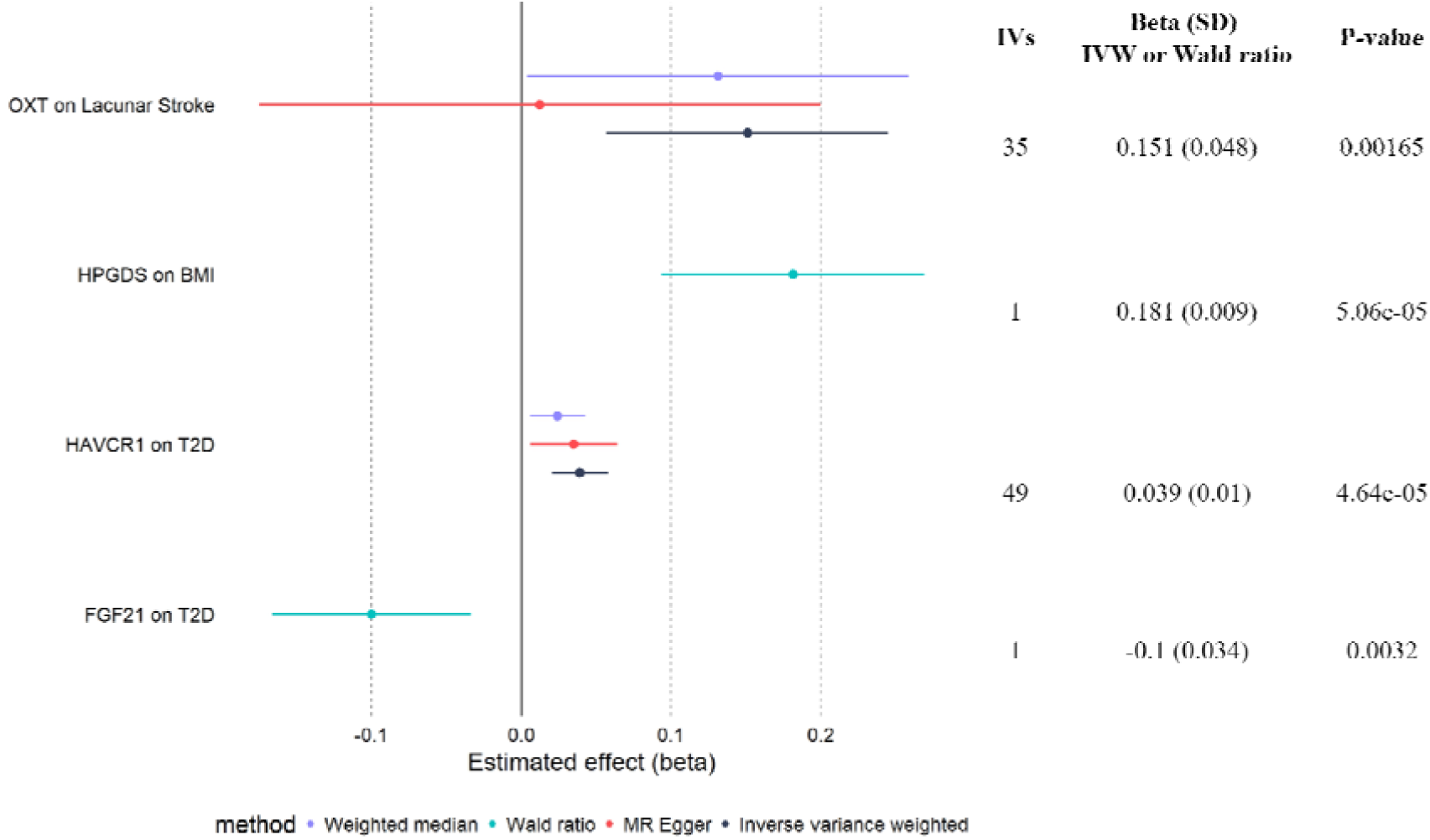
Forest plot representing the effect sizes of two-sample MR for the four significant proteins-traits. The IVW method along with sensitivity analyses are shown for proteins with more than one IV, and the Wald ratio estimates are shown for proteins with only one IV.

## 4. Discussion

In the present study, we performed plasma metabolomic and proteomic profiling for the first time on a group of individuals who follow a unique dietary pattern, and have revealed extensive metabolic reprogramming associated with abstinence from meat, fish, dairy products and eggs. This dietary behaviour resembles an interventional study, but in this case the intervention is practiced voluntarily for religious reasons, and results in overall lower intake of protein, but not of energy [20, 21].

Animal product restriction-associated changes in metabolite levels resulted in profiles compatible with a positive impact on health. In addition to reductions in levels of multiple lipid classes, we found reductions in mTORC1 agonists valine and leucine. mTORC1 integrates nutrient and hormonal cues to regulate cellular processes linked to growth and proliferation, and ultimately healthspan and longevity [2, 3]. Methionine restriction also downregulates mTORC1 activity, driving increased levels of FGF21 and improved metabolic profiles [2, 34, 42]. Given that animal products are rich in methionine, and also that vegetarian and vegan individuals typically displaying lower plasma levels of this amino acid [7, 63, 64], we suggest that the observed increase in FGF21 levels may also be driven by reduced intake of methionine. Overall, changes in protein levels associated with dietary restriction were in a direction suggesting at least some favourable effects on health, further confirmed through MR for FGF21, HAVCR1 and HPGDS.

In addition to positive effects on health we also find effects that are potentially negative. Increased levels of FGF21, but also of restriction-associated changes in levels of OXT, SPP1, FGF23, PTH1R and PTHrP, may exert detrimental effects on bone homeostasis [44], [50], [65]. We have also revealed an effect of OXT on increased risk for lacunar stroke. Increased risk of stroke in vegetarians has been reported previously [6], but the mechanisms underlying this observation are not clear. Proposed explanations include low levels of LDL cholesterol, high levels of homocysteine due to low vitamin B12, and low intake of animal protein [6], but further investigation is required to understand how risk is determined. Furthermore, due to their role in regulating dietary preference, impaired signalling through FGF21 and OXT underlies dietary choices that exacerbate obesity and linked diseases [66]. Understanding the signalling pathways driven by these hormones will aid in harnessing the mechanisms that govern feeding regulation and will contribute to the development of development of FGF21 and OXT-based pharmacotherapies that promote healthy diet choice.

Our work has highlighted high-value candidates to evaluate for their potential therapeutic effects and findings from the present study could potentially inform the development of therapeutics that mimic molecular responses that promote healthy aging and have a beneficial impact on health. In addition to the potential of restriction-associated proteins as pharmaceuticals, we have revealed changes in the abundance of potentially druggable proteins suggesting that dietary interventions (**Figure 4, Supplementary Figure S9, Table 1**) can impact the kinetics of drug-target binding [67] and may be used to fine-tune the action of drugs.

Although the molecular changes we have uncovered are extensive, they are most likely transient, especially for metabolite profiles which were identical between dietary groups when both were on an omnivorous diet. On the other hand, we detected consistent differences in protein levels across dietary groups, suggesting at least some long-term molecular effects at the protein level. In addition to ALPP, such proteins include CXCL17, a cytokine linked to development of obesity and metabolic dysfunction [68], LAMP3, a regulator of hepatic lipid metabolism [69], and OMD, a regulator of osteogenesis [70]. Our study design has also enabled us to uncover changes in protein abundance that are likely seasonal, and understudied in humans despite the known seasonal trends of various aging-related phenotypes including cardiovascular disease, allergies, autoimmune conditions, and psychiatric disorders [71].

This work is limited by the absence of dietary intake data for study participants. However, given the very specific guidelines outlined by the Greek Orthodox Church, the dietary pattern under study is highly consistent between different groups of practicing individuals. Indeed, for overlapping measurements, our findings are in line with those from two studies covering the same restriction period (Lent) [20, 21]. Secondly, profiling was not performed immediately prior to initiation of animal product restriction. However, given that the dietary patterns preceding both T1 and T2 are very similar [10], we expect that measurements prior to initiation of dietary restriction at T2 would have been very similar to measurements at T1. Finally, molecular levels assessed were in plasma samples, thus representing the biology of multiple tissues and not enabling us to ascertain tissue-specific mechanisms.

In summary, our results suggest that animal product restriction results in rapid and extensive molecular reprogramming with mostly positive, but some likely detrimental effects on health. Further work is required to better understand how underlying molecular mechanisms including changes in gene transcription, in immune cell type composition and in the gut microbiome, affect mechanisms linked to metabolic health, aging and risk for disease. Furthermore, longitudinal studies are required to map long-term effects on health and to evaluate whether simple dietary supplementation (e.g. with vitamin B12) can minimise negative consequences.

## Data Availability

All data produced in the present study are available upon reasonable request to the authors. Proteomics and metabolomics summary statistics are available at Zenodo.

https://zenodo.org/records/10985423

## Abbreviations

T1: Timepoint 1
T2: Timepoint 2
BMI: Body mass index
LDL: low-density lipoprotein
HDL: high-density lipoprotein
PR: periodically restricted
NR: non-restricted
SBP: systolic blood pressure
DBP: diastolic blood pressure
NMR: nuclear magnetic resonance
QC: quality control
PCA: principal component analysis
FDR: false discovery ratio
EFO: experimental factor ontology
MR: mendelian randomization
T2D: type 2 diabetes
BCAAs: branched chain amino acids
FGF21: Fibroblast Growth Factor 21
FOLR2: Folate Receptor Beta
SUMF2: Sulfatase Modifying Factor 2
HAVCR1: Hepatitis A Virus Cellular Receptor 1
PLA2G1B: Phospholipase A2 Group IB
OXT: Oxytocin
HPGDS: Hematopoietic Prostaglandin D Synthase
SPP1: Secreted Phosphoprotein 1

## CRediT authorship contribution statement

Conceptualization: KR, OB and ASD. Methodology: KR, OB, PM, A. Skoulakis and ASD. Formal analysis: OB, A. Simistiras, CE, PM, A. Skoulakis, and YCP. Investigation: KR, AD, SG and ASD. Data curation/software: OB, A. Simistiras, CE, PM, A. Skoulakis, and YCP. Visualization: OB, A. Simistiras, CE, PM, A. Skoulakis. Supervision: EZ and ASD. Writing- Original draft: OB and ASD. Writing-review and editing: KR, OB, A. Simistiras, CE, PM, A. Skoulakis, YCP, AD, SG, GK, EZ and ASD. Project administration: ASD. Funding acquisition: ASD.

## Acknowledgements

The authors are grateful to the FastBio study participants and to the Interbalkan Hospital Staff. We would also like to thank Dr Dimitrios Rouskas, Dr Pavlos Rouskas and Dr Loukas Kipouros for their invaluable help with sample collection. We acknowledge the technical support of Core Facility Metabolomics and Proteomics at Helmholtz Munich and thank Dr Stefanie Hauck and Dr Agnese Petrera for help with proteomics assays. We would also like to thank Professor Iannis Talianidis for his very helpful comments and input throughout the course of this project and Dr Aliki Perdikari for useful discussions.

## APPENDIX A. Supplementary Information

## Data availability

Metabolomics and proteomics data have been deposited in ref [72].

