## Supplement for "Periodic dietary restriction of animal products induces metabolic reprogramming in humans with effects on health"

**Supplementary Information**

**Supplementary Text S1. FastBio study inclusion and exclusion criteria**

Inclusion criteria:

- Healthy female and male subjects 18 – 75 years of age at the time of enrolment. One participant turned 76 between time of enrolment and first sampling timepoint (T1).
- Able to provide signed and dated informed consent. Willing to provide blood samples.
- Individuals who had practiced periodic animal product restriction for at least ten years (for the PR group).
- Individuals who had not practiced any kind of specific diet including veganism, vegetarianism, caloric restriction, intermittent fasting (for the NR group).

Exclusion criteria:

- Use of antibiotic, antifungal, antiviral or antiparasitic drugs six months prior to T1.
- Acute disease, defined as the presence of a moderate or severe illness with or without fever, at sampling timepoints.
- Alcohol or drug abuse two years prior to T1.
- Participants who are normally periodically abstaining from animal products, but had to alter their diet for a specific reason (e.g. pregnancy).

**Supplementary Text S2. Dietary pattern of periodic animal product restriction specified by the Greek Orthodox Church**

Individuals following the dietary regimen of the Greek Orthodox Church practice restriction of animal products through abstinence from meat, fish, dairy products and eggs for 180-200 days annually. Consumption of shellfish and molluscs is permitted on all days of restriction. Restriction is practiced on Wednesdays and Fridays throughout the year (excluding the week immediately after Christmas, Easter and the Pentecost) and over four extended periods annually as follows (**Figure 1b**):

- 40 days before Christmas (abstinence from meat, dairy products and eggs, consumption of fish is permitted except on Wednesdays and Fridays).
- 48 days before Easter (Lent) (abstinence from meat, fish, dairy products and eggs, consumption of fish is permitted only on March 25^th^ and on Palm Sunday).
- 0-30 days in June (abstinence from meat, dairy products and eggs, consumption of fish is permitted).
- 15 days before August 15th (Assumption) (abstinence from meat, fish, dairy products and eggs, consumption of fish permitted only on August 6^th^).

**Supplementary Text S3. Comparison of metabolite and protein profiles between dietary groups at T1 and T2**

At T1, when both groups were on an omnivorous diet, no differences in metabolites levels were detected **(Supplementary Figure S4, Supplementary Table S2**). At T2, following restriction for the PR group, we report 102 found at significantly differences abundance between dietary groups. Of these, 97 (95%) were also detected in the PR group upon from T1 to T2. Furthermore, at T1 we find a similar mortality score between PR and NR groups (mean score 0.11 in PR vs 0.079 in NR, p=0.951 (**Supplementary Figure** **S5**), but at T2 we report significantly lower values for PR individuals (mean score -0.91 in PR vs 0.70 in NR, p=0.007). When comparing the association patterns from T2 with complex diseases, the trends we found were similar to the association profiles comparing the two timepoints in PR individuals (**Supplementary Figure S4**). Similarly, we examined differences in protein levels between dietary groups and found that 18 and 60 proteins existed at significantly different levels at T1 and T2 respectively (**Supplementary Figure S7**). Of these, 10 were detected at both timepoints likely reflecting long-term differences between dietary groups. Unique associations at T2 were over six times as many compared to T1 (50 vs 8), reflecting acute effects of animal product restriction, with FGF21 being the most significant finding. Of the 50 proteins that were unique to T2, 31 (62%) were also detected as uniquely associated in PR individuals from T1 to T2 supporting the idea of effects linked to animal product restriction.

**Supplementary Text S4. Integration of metabolite and protein datasets**

To explore links between metabolites and proteins we performed unsupervised correlation analysis using the sPLS (Sparse Partial Least Squares) method of the mixOmics package [1, 2]. This method identifies linear combinations of the variables from each omics dataset and reduces the dimensionality, while also performing variable selection through LASSO penalization. To enhance power, we analysed all participants at both timepoints, accounting for repeated measures and used variable selection to focus on a subset of optimally selected key predictor metabolites and proteins. For the selection of the number of variables per component per dataset, 10-fold cross-validation was repeated five times and the outcome that maximized the correlation between the predicted and the actual components was chosen. Optimal selection involved two components with five variables each. The first component captured animal product restriction-associated effects, with five PR-unique differentially abundant proteins (HAVCR1, ESM1, SPP1, SPON2, FGF21) displaying correlations with IDL particles (**Supplementary Figure S8**). Although we do not have evidence for causality, we suggest that IDL particles which were affected by dietary restriction, may comprise a promising metabolite type to investigate further for effects on health. The second component captured effects not linked to animal product restriction with five proteins (LDLR, NPY, AGRP, CD38, MFGE8) correlating positively with L and XL VLDL particles and with triglycerides (**Supplementary Figure S8**). Studies in rats have shown that NPY controls hepatic secretion of VLDL triglycerides [3], suggesting that our work can also be used to understand links between metabolites and proteins independent of dietary restriction.

**Supplementary Text S5. Over-representation analysis**

The Olink Explore 1536 panel comprises four separate panels that are enriched for proteins with functions in specific biological processes: cardiometabolic, inflammation, oncology, neurology. Pathway over-representation analysis was performed for differentially abundant proteins using the online tool ShinyGO v.0.77 [4] and applying the total number of proteins tested (N=1,455) as background (Olink 1536 panel proteins after QC). KEGG (Release 86.1) was used as the pathway database. KEGG pathways with FDR adjusted p-value ≤ 0.05 were considered to be over-represented. We conducted over-representation analysis to test if proteins detected at altered levels were enriched for specific biological pathways. For each comparison, we tested total differentially abundant proteins for over-represented pathways, but also explored whether there were over-represented pathways when grouping proteins as up or down regulated.

Using the tested 1,455 proteins as a background, over-represented pathways were detected for:

- PR unique downregulated (N=102): metabolic pathways.
- NR unique downregulated (N=24): human papillomavirus infection, bladder cancer.
- T1 unique total (N=50): PI3K-Akt signalling pathway, focal adhesion, relaxin signalling pathway, ECM-receptor interaction, AGE-RAGE signalling pathway in diabetic complications.
- T1 unique downregulated (N=5): PI3K-Akt signalling pathway, relaxin signalling pathway, AGE-RAGE signalling pathway in diabetic complications, pathways in cancer, Ras signalling pathway, focal adhesion.

Given that the proteins assayed in the present study derive from a panel that is pre-enriched for specific functions, our analysis is underpowered and biased towards false negatives. Similarly, when applying all protein coding genes as background (19,566 HGNC genes) to interrogate over-representation of pathways, our results are biased for false positives. Therefore, this analysis is likely underpowered to provide substantial insight for the present study.

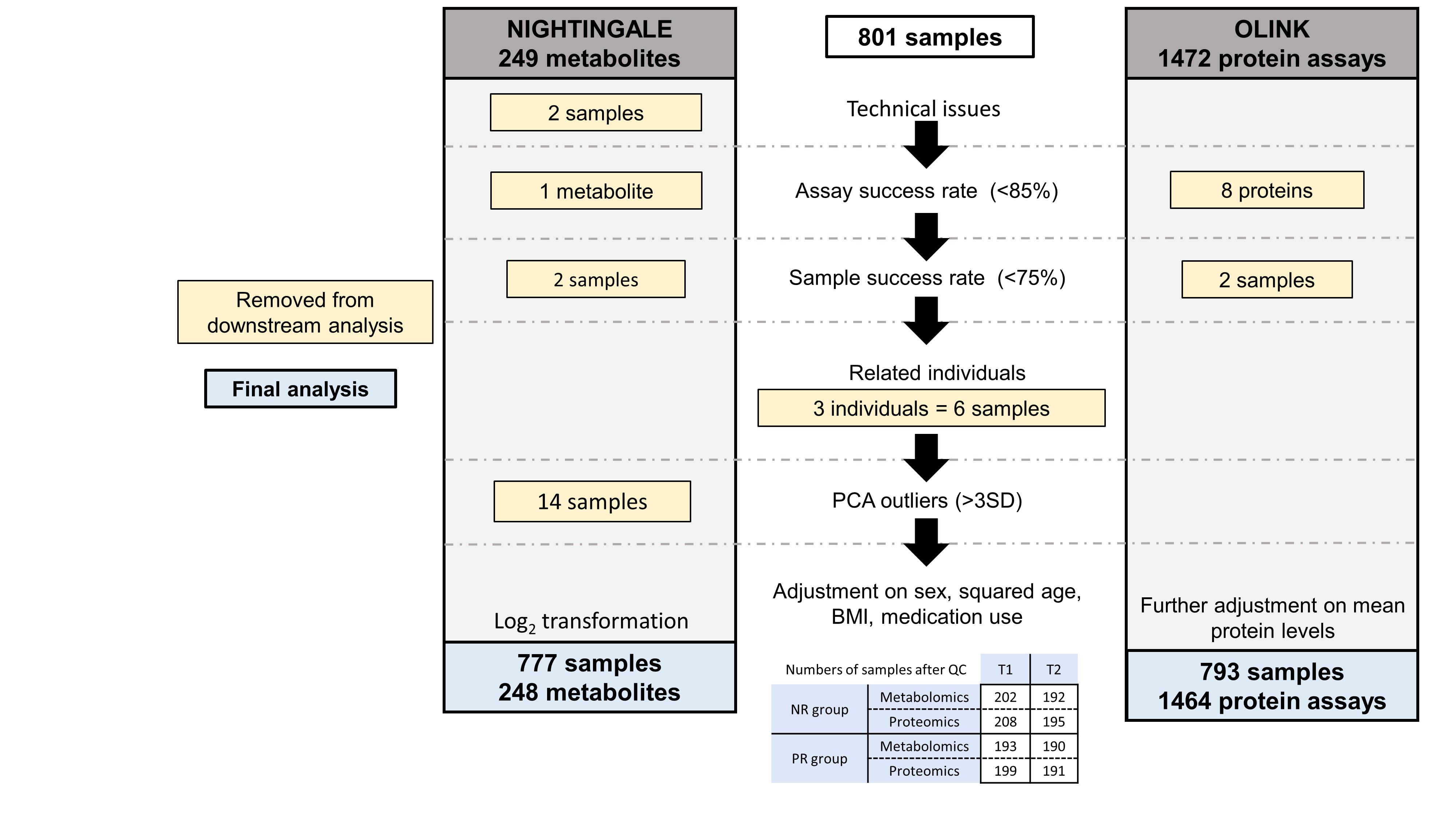

**Supplementary Figure S1 Quality control flowchart for metabolomic and proteomic data.** This schema represents the quality control performed, showing numbers of metabolites and proteins, as well as samples removed at each step. Final numbers of metabolites and proteins and of samples in each dietary group and at each timepoint are shown in the bottom table.

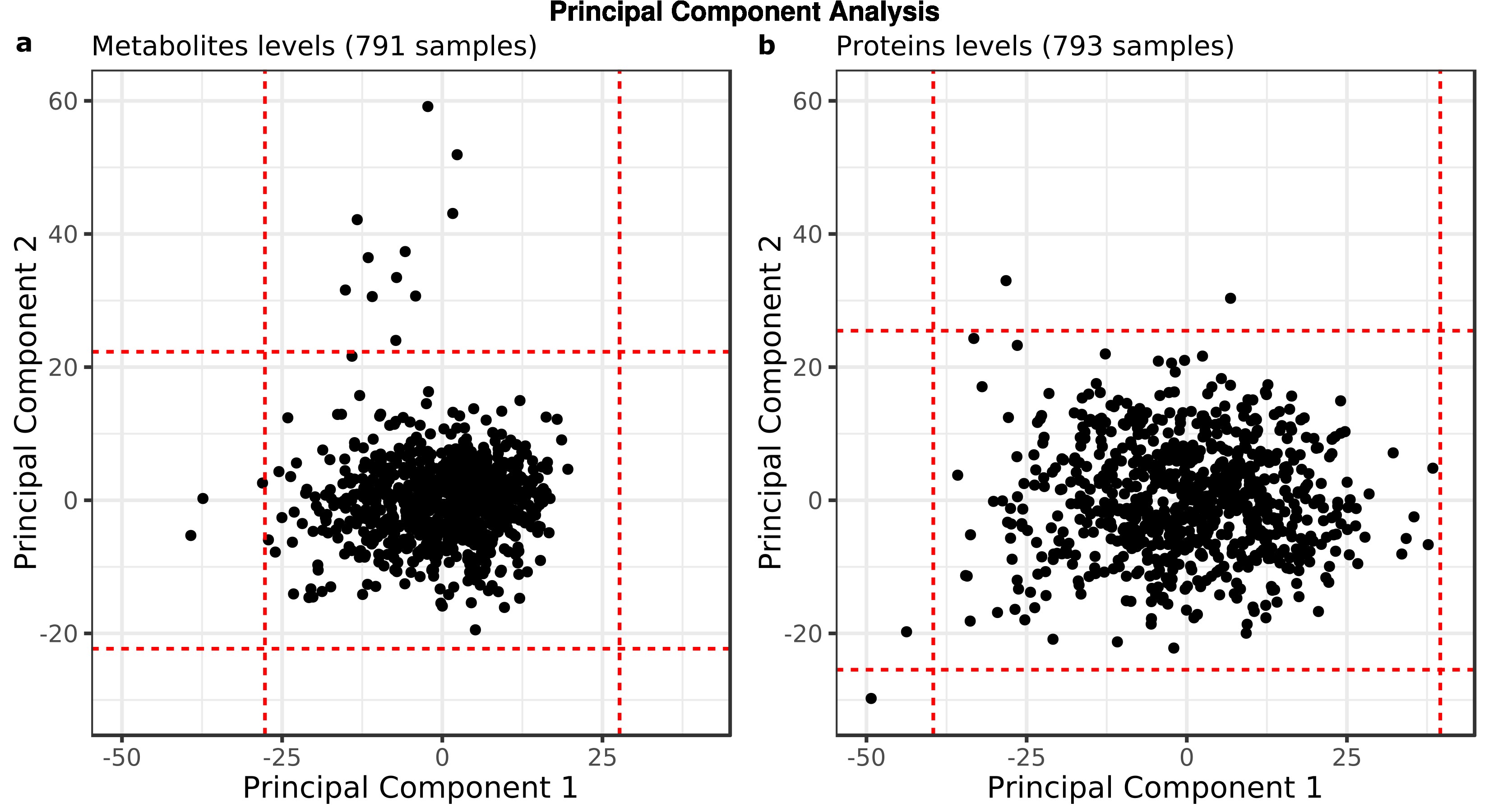

**Supplementary Figure S2 Principal Component Analysis (PCA) of metabolite (a) and protein (b) levels.** PCA was performed on metabolite and protein levels before data transformation. Red lines correspond to +/- 3 standard deviations from the mean.

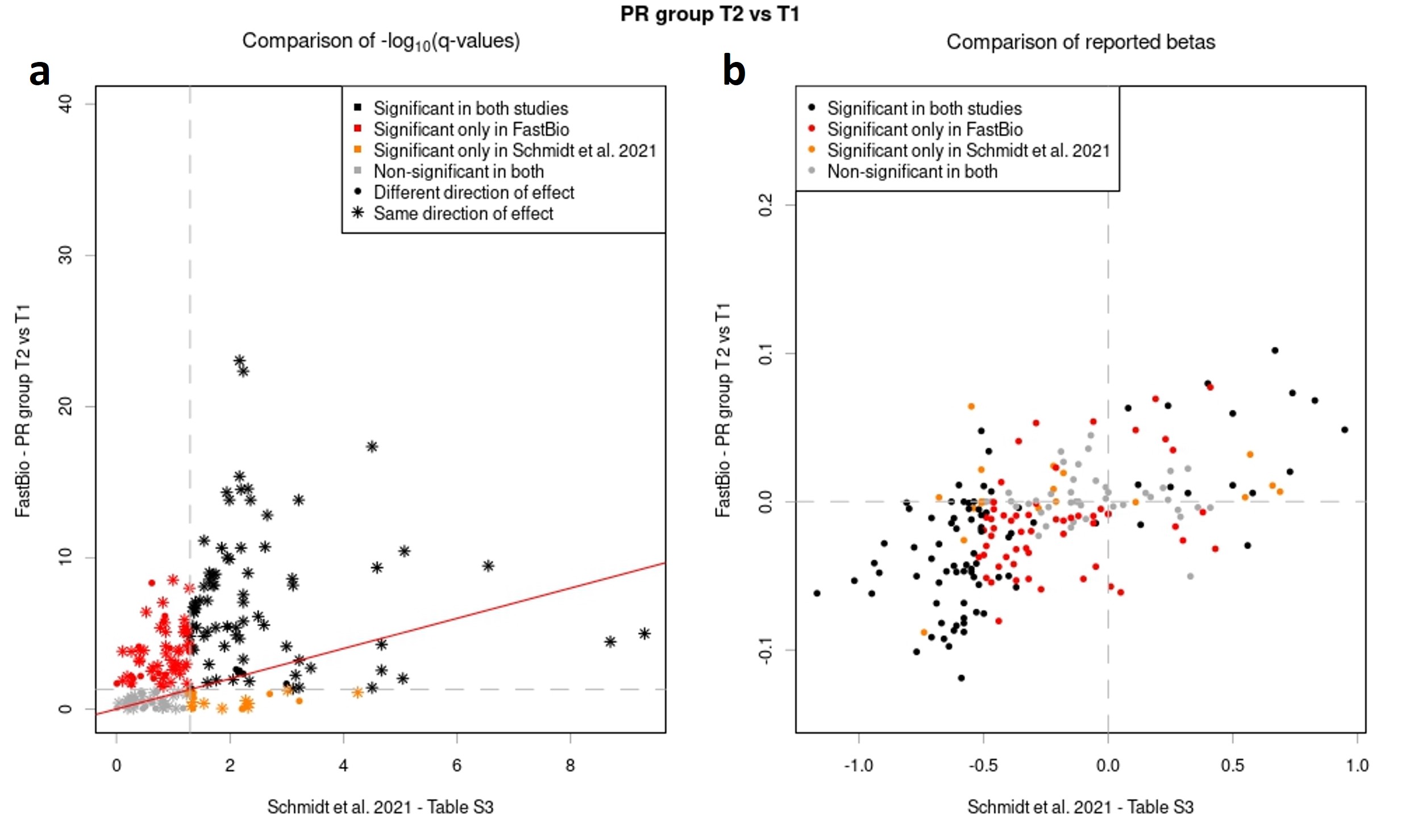

**Supplementary Figure S3 Comparison of differentially abundant metabolites detected from T1 to T2 in the PR group, to metabolites with significantly different levels detected between vegan and meat-eating individuals.** Associations between metabolite profiles and timepoints in the PR group are compared to associations from [5], both in terms of p-values (panel **a**) and effect sizes (panel **b**). Metabolites are coloured according to their significance in the two studies. Star-shaped points in panel **a** represent a concordant direction of effect.

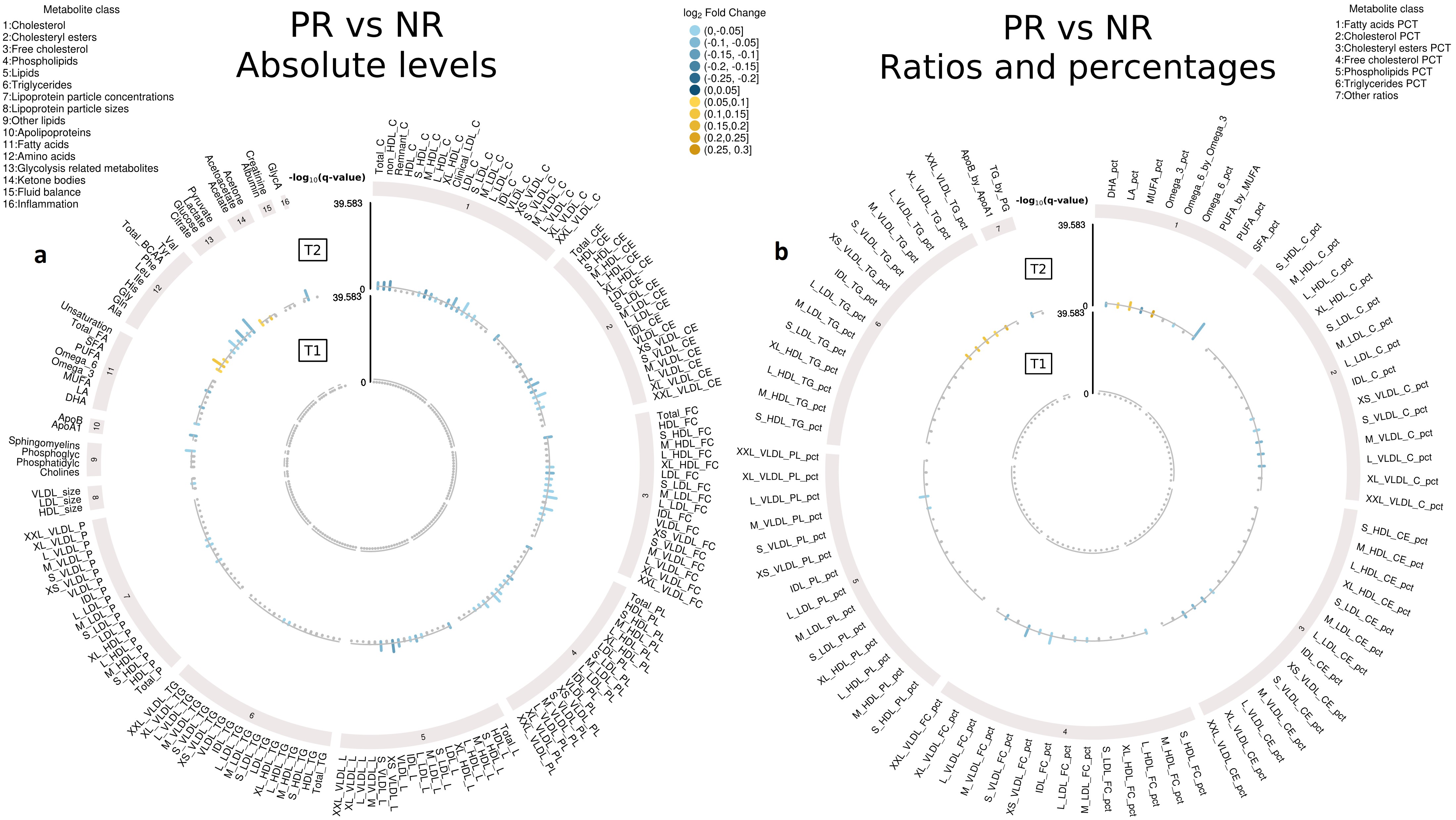

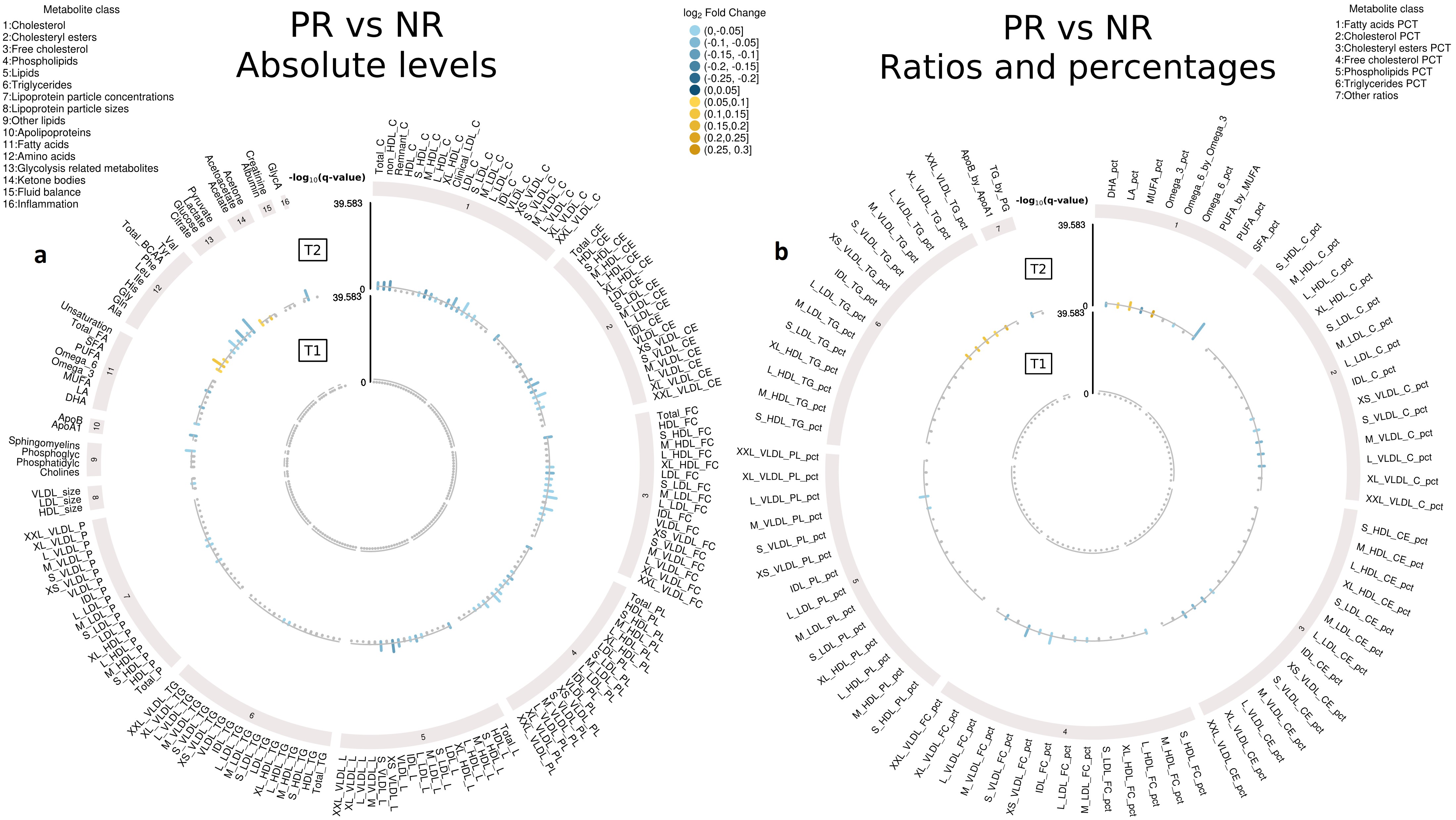

**Supplementary Figure S4 Differentially abundant metabolites detected between dietary groups at T1 and at T2.** Differences in metabolite profiles between dietary groups are shown in the outer circle for T2 and in the inner circle for T1 (absolute values in panel **a**, ratios and percentages in panel **b**,). The -log_10_ of the FDR-adjusted p-value (q-value) is represented in the y-axis. Yellow bars represent higher levels in the PR group whereas blue bars represent lower levels in the PR group at each timepoint. Metabolites shown in grey are not significant.

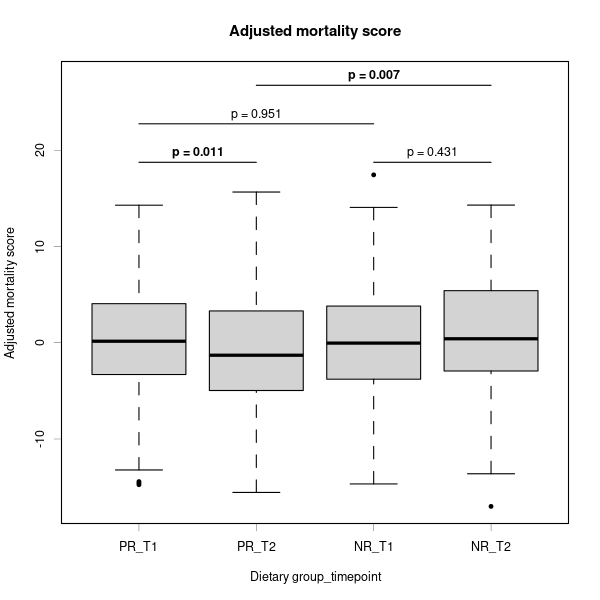

**Supplementary Figure S5 All-cause mortality score for each dietary group at each timepoint.** The distributions of the 14-metabolite mortality score developed in ref. [6] are represented as boxplots and significant association p-values from a regression model of the scores against timepoints and dietary groups are shown in bold.

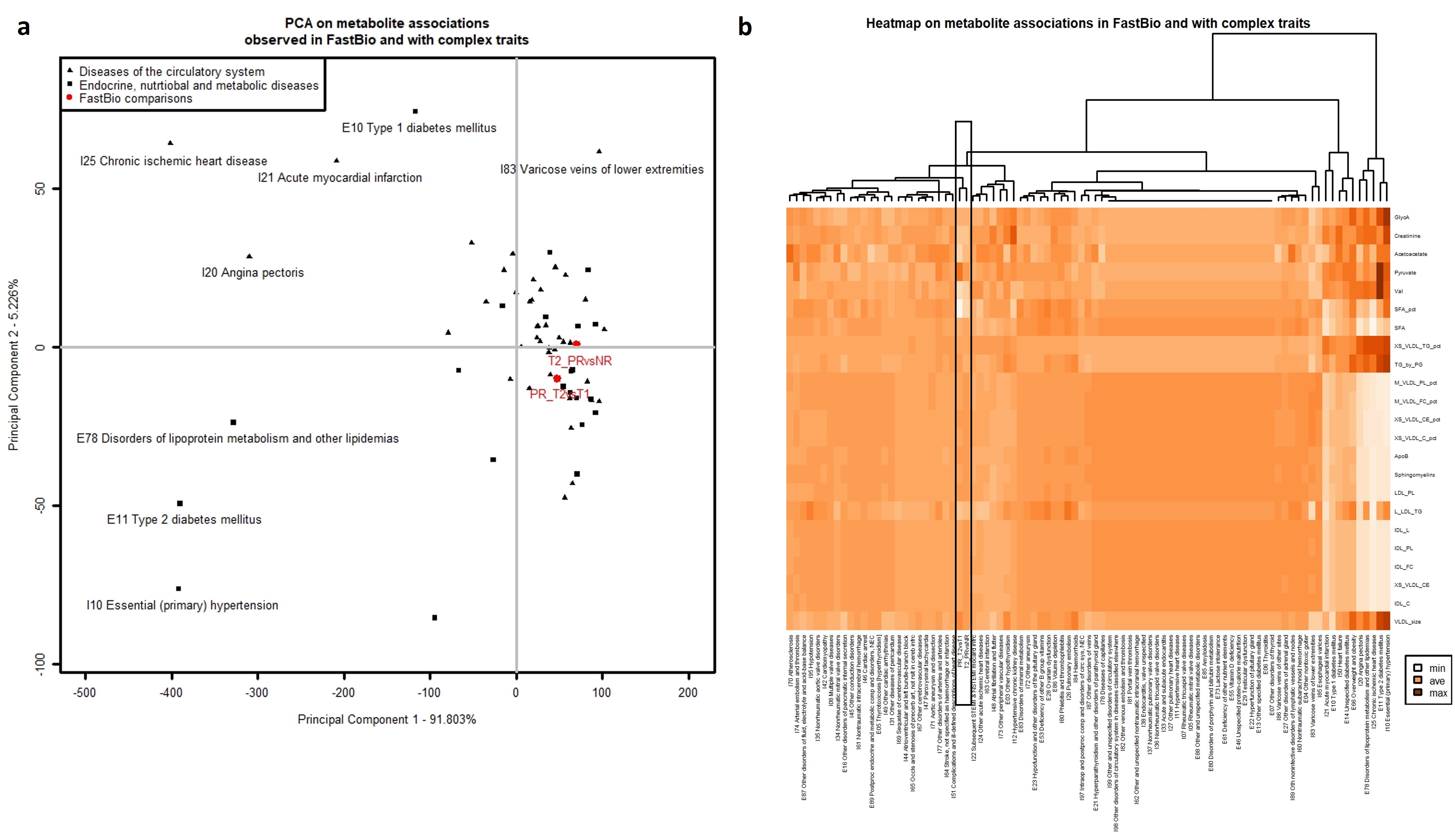

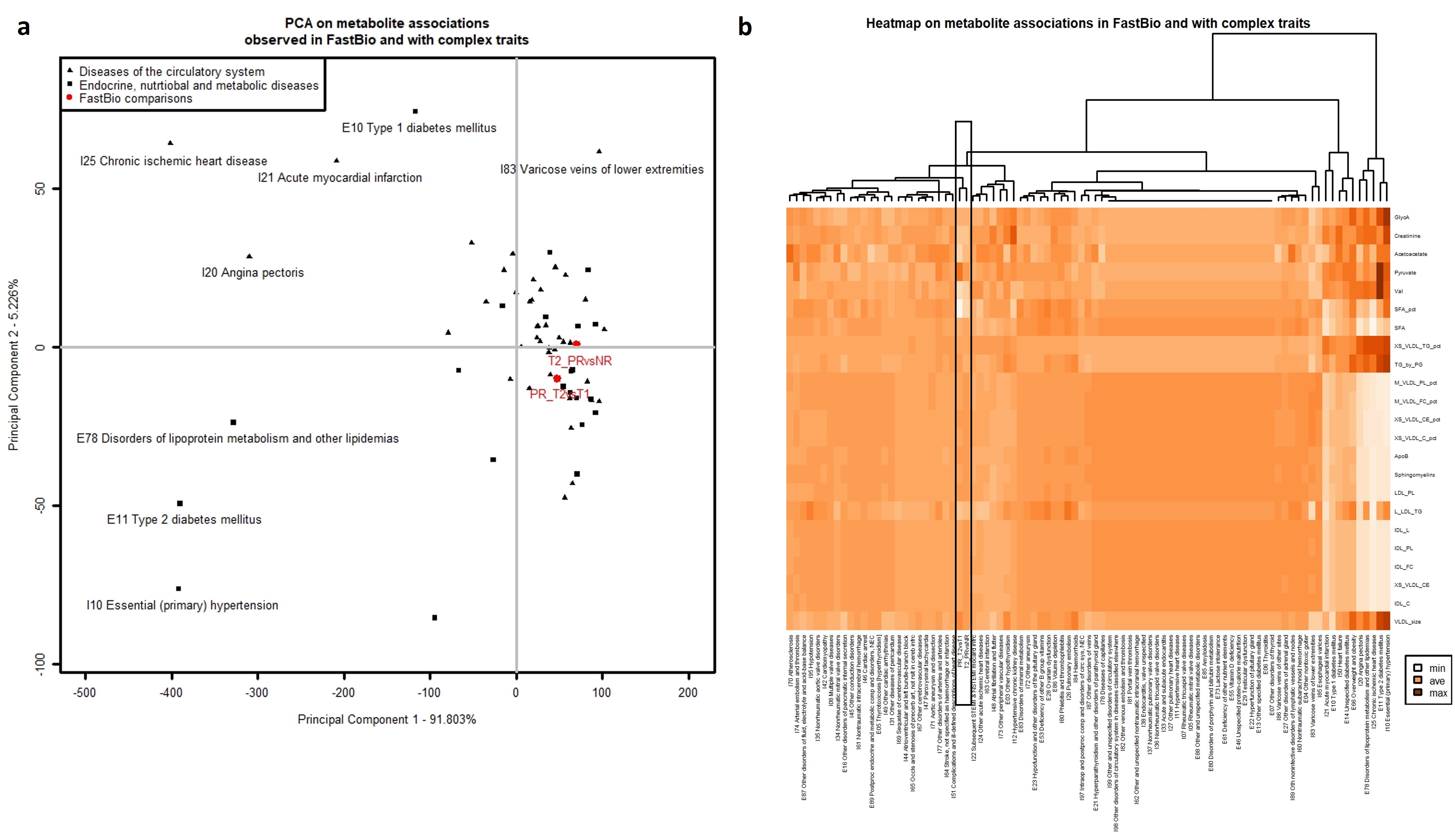
**Supplementary Figure S6 Principal component analysis (PCA) and heatmap of metabolite associations with complex diseases and in FastBio comparisons. a,** PCA constructed on the metabolite associations (t-statistics) described in the PR group (T2 vs T1) and at T2 (PR vs NR), and reported with “diseases of the circulatory system" and “endocrine, nutritional and metabolic diseases” in the UK Biobank (ref. [7]). **b,** Heatmap of the same associations where the most significant metabolite for each metabolite class has been selected based on the T2 vs T1 comparison in the PR group. PR and T2 comparisons are highlighted in this heatmap. In the PCA, the two first principal components are represented, and labels are reported for PR and T2 comparisons and for the diseases showing the most distinguishable pattern of associations. The PCA is mainly constructed on associations with complex diseases and driven by those showing the strongest associations with metabolites. From the heatmap, this corresponds to type 2 diabetes, chronic ischemic heart disease, essential hypertension, angina pectoris and disorders of lipoprotein metabolism which are strongly associated with lipoproteins. Other diseases, as well as PR and T2 associations, are close on the PCA as they show much lower magnitudes of associations with these metabolites. We can therefore interpret comparisons within the PR group and at T2 with diseases the furthest away, for which animal product restriction seems to be protective. In the dendrogram, the PR and T1 comparisons are grouped together in a single cluster distinct from the other diseases, highlighting an overall specific pattern of associations different from the selected complex diseases.

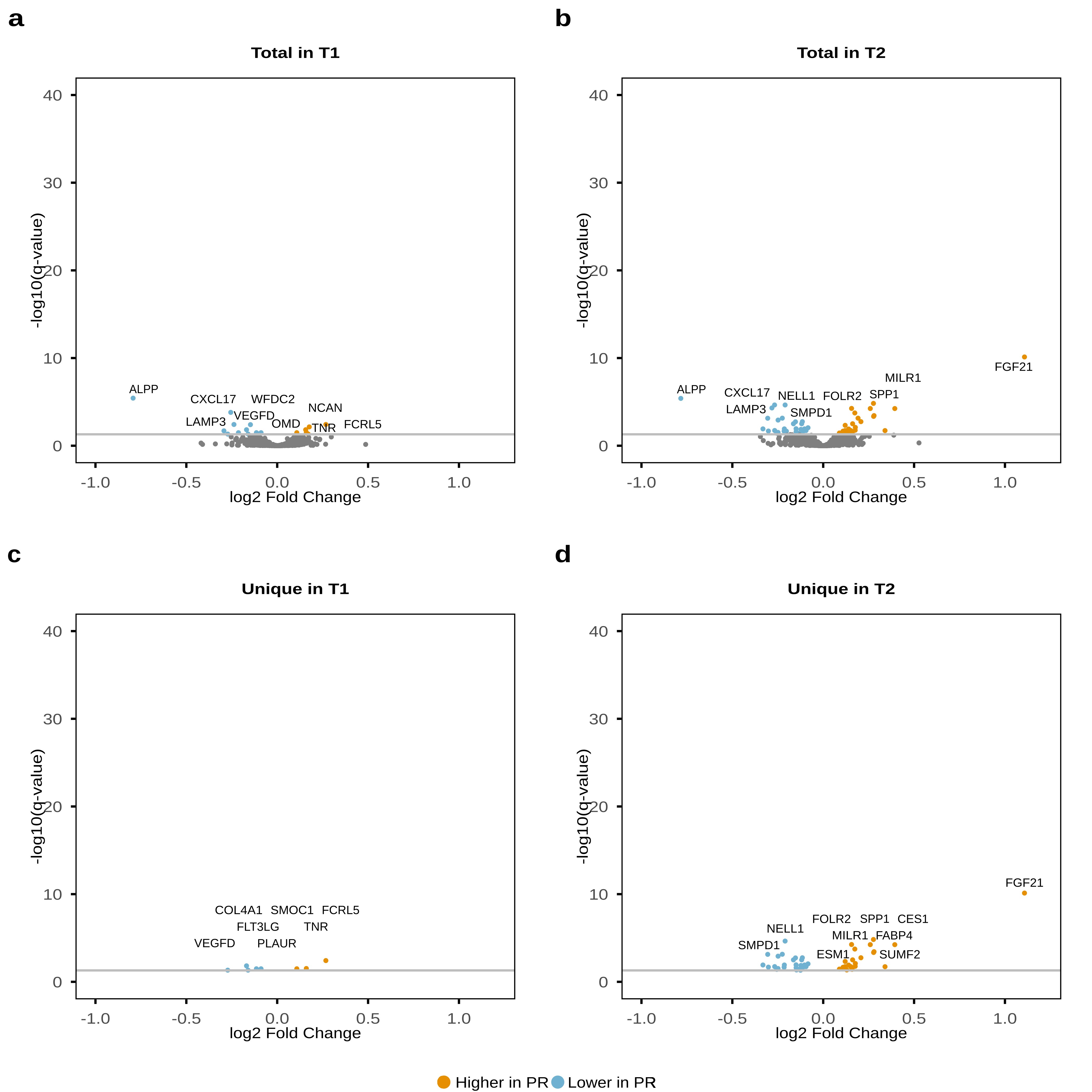

**Supplementary Figure S7 Differentially abundant proteins detected between dietary groups at T1 and at T2.** Total differentially abundant proteins detected at T1 (**a**) and at T2 (**b**). Unique differentially abundant proteins detected at T1 (**c**) and at T2 (**d**). Differentially abundant proteins found in lower levels in the PR group are shown in blue whereas differentially abundant proteins found in higher levels in the PR group are shown in yellow. Proteins shown in grey are not significant.

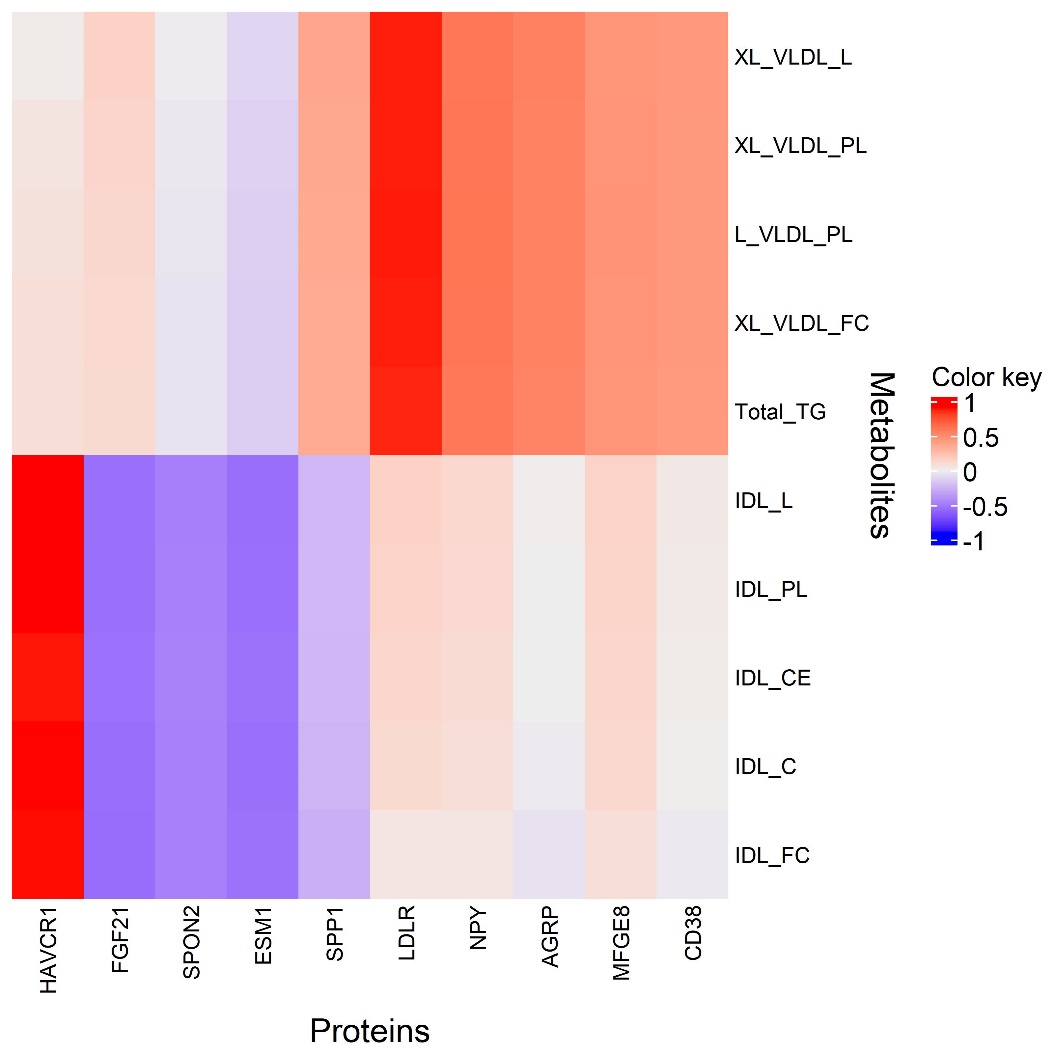

**Supplementary Figure S8** **Metabolite-protein correlations.** Correlations between optimally selected proteins (x axis) and metabolites (y axis) by the sPLS method in mixOmics.

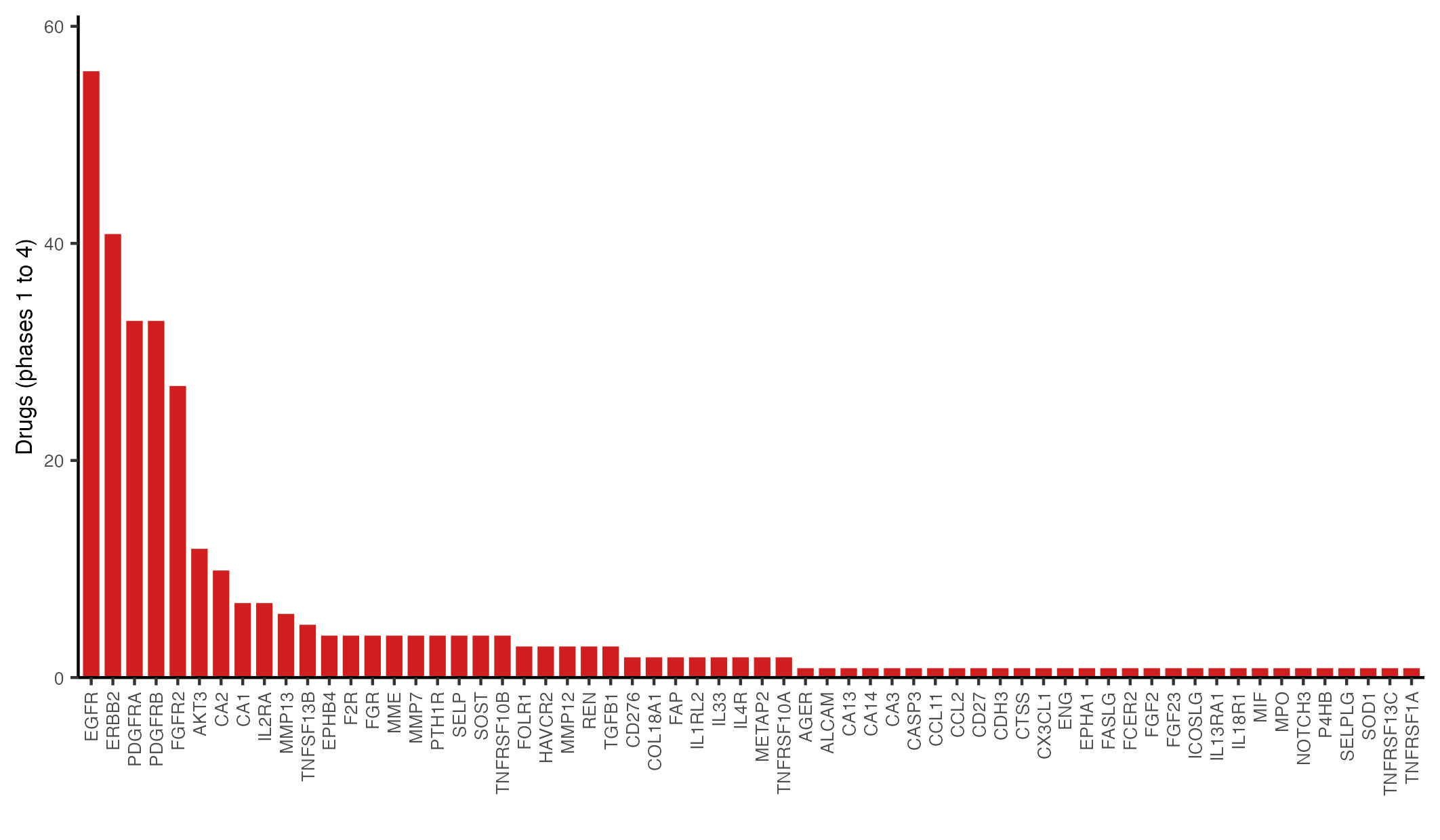

**Supplementary Figure S9 PR-unique differentially abundant proteins as targets of phase 1-4 drugs.** Number of phase 1-4 drugs (excluding clinical trials with withdrawn, unknown or terminated status) that target differentially abundant proteins detected only in PR individuals.

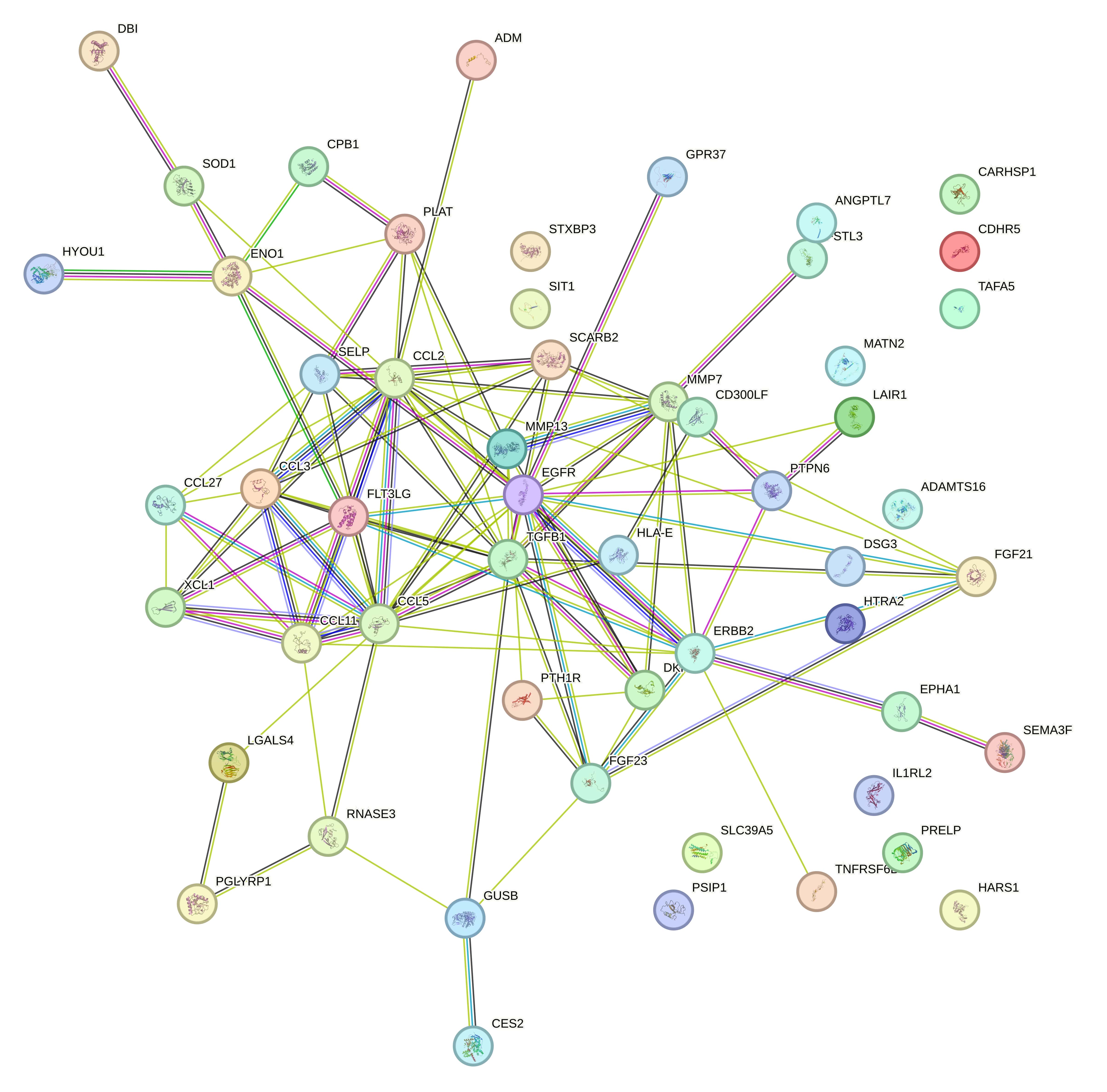

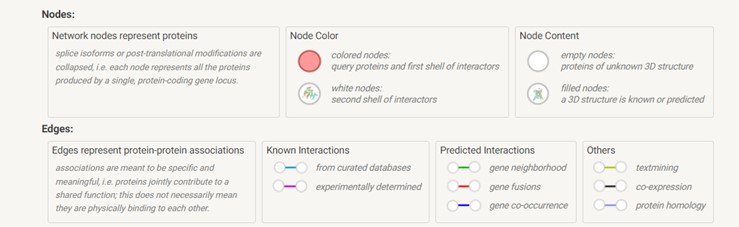

**Supplementary Figure S10.** **Interaction network of 53 proteins with suggestive links to FGF21 signalling in humans.** The nodes represent proteins, and edges represent protein-protein interactions.

**Supplementary Table S1. Mendelian randomization analysis.** (xlsx)

**Supplementary Table S2. Overview of differentially abundant metabolites and differentially abundant proteins between timepoints and dietary groups.** (xlsx)

**Supplementary Table S3. Phase 1-4 drugs targeting PR-unique proteins and associated conditions.** (xlsx)

**Supplementary Table S4. Phase 1-4 clinical trials for proteins displaying the greatest magnitude of animal product restriction-associated changes.** (xlsx)

**Supplementary Table S5. Proteins with suggestive links to FGF21**. Upon adjustment for FGF21 levels, 53 out of the 409 PR-detected differentially abundant proteins were no longer significant. (xlsx)
